# A feasibility study to test a novel approach to dietary weight loss with a focus on assisting informed decision making in food selection

**DOI:** 10.1101/2021.05.21.21257479

**Authors:** Mindy H. Lee, Catherine C. Applegate, Annabelle L. Shaffer, Abrar Emamaddin, John W. Erdman, Manabu T. Nakamura

## Abstract

Obesity is a significant contributor to the development of chronic diseases, some of which can be prevented or reversed by weight loss. However, dietary weight loss programs have shortcomings in the success rate, magnitude, or sustainability of weight loss. The Individualized Diet Improvement Program’s (iDip) objective was to test the feasibility of a novel approach that helps individuals self-select a sustainable diet for weight loss and maintenance instead of providing weight loss products or rigid diet instructions to follow.

The iDip study consisted of 22 dietary improvement sessions over 12 months with six months of follow-up. Daily weights were collected, and a chart summarizing progress was provided weekly. Six 24-hour dietary records were collected, and dietary feedback was provided in the form of a protein-fiber plot, in which protein/energy and fiber/energy of foods were plotted two-dimensionally together with a target box specific to weight loss or maintenance. An exit survey was conducted at 12 months.

Twelve (nine female, 46.3±3.1 years (mean±SE)) of the initial 14 participants (BMI>28 kg/m^2^) completed all sessions. Mean percent weight loss (n=12) at six and 12 months was -4.9%±1.1 (p=0.001) and -5.4%±1.7 (p=0.007), respectively. Weight loss varied among individuals at 12 months; top and bottom halves (n=6 each) achieved - 9.7%±1.7 (p=0.0008) and -1.0%±1.4 weight loss, respectively. The 24-hour records showed a significant increase in protein density from baseline to final (4.1g/100kcal±0.3 vs. 5.7g/100kcal±0.5; p=0.008). Although mean fiber density showed no significant change from the first month (1.3g/100kcal±0.1), the top half had significantly higher fiber/energy intake than the bottom half group. The survey suggested that all participants valued the program and its self-guided diet approach.

In conclusion, half of the participants successfully lost >5% and maintained the lost weight for 12 months without strict diet instructions, showing the feasibility of the informed decision-making approach.

## Introduction

With the advancement of food production globally, obesity has become a worldwide epidemic, increasing the risk of morbidity and mortality [1, 2]. In the United States, 70.2% of adults are obese (body mass index, BMI>30 kg/m^2^) or overweight (BMI>25 kg/m^2^), with 7.7% having extreme obesity (BMI>40 kg/m^2^) [3]. Overweight and obesity are risk factors for several comorbidities, including type 2 diabetes mellitus, cardiovascular disease, and cancers. Based on 2013 US medical spending, obesity and its complications cost $342.2 billion annually (28.2% of all medical expenditures) [4]. Modest weight losses of 3-5% initial body weight are capable of lowering blood triglycerides, fasting blood glucose, and improving glycemic control. Further weight losses enhance these benefits, and can also reduce blood pressure, cholesterol, and need for medications [2, 5].

Currently available weight loss methods do not reliably achieve medically significant weight loss as they are insufficient in success rate, sustainability, or magnitude of weight loss. Exclusion of certain food groups is a common approach of popular dieting programs, which can result in some weight loss, but high attrition rates often result due to a lack of dietary adherence [6-8]. For example, commonly used dieting programs, including Atkins (carbohydrate restriction), Zone (substituting carbohydrate with protein), and Ornish (reduction of animal foods), can induce modest weight losses of <5 kg but lack a high success rate and long-term sustainability [9-12]. Participants grow fatigued with the strict recipes and are unable to adapt the plan to their daily lives [13, 14]. This translates to low adherence, unsuccessful weight loss, and lack of diet sustainability.

A negative energy balance is a requirement for weight loss. Calorie monitoring is often proposed to achieve this and is recommended by health professionals [15, 16]. However, it is laborious and frequently inaccurate [17, 18]. Often, programs utilizing calorie counting or food journals lead to high attrition and thus, insignificant long-term weight loss [19-21]. More importantly, the protein requirement increases during periods of negative energy balance compared with the requirement during weight maintenance [22, 23]. Focusing only on energy reduction may result in exacerbating the loss of lean body mass and decreased efficiency in weight loss [23].

Successful, clinically significant weight losses can be achieved using meal replacement products; however, weight maintenance remains a substantial challenge for this approach [5]. Very-low-calorie diets (VLCD) in the form of meal replacements have shown clinically significant weight loss in the short term, but VLCD participants are less successful in weight maintenance compared to a restricted-energy whole-food diet [24-26]. Even when a VLCD was combined with intensive dietary and lifestyle intervention programs using counseling, education, and exercise, participants regained most or all of the lost weight during the follow-up periods [5, 27].

Pharmaceuticals are available for treating obesity, such as orlistat and phentermine/topiramate, and work by reducing fat absorption and suppressing appetite, respectively. However, these medications result in only 3-6% weight loss and can include several side effects such as reduced fat-soluble vitamin absorption or gastrointestinal upset [28]. As a result of the low efficacy in medicine and lifestyle interventions, bariatric surgery remains the most reliable weight-loss treatment and can reduce obesity complications, such as hyperglycemia [29]. Bariatric surgery does have significant drawbacks, including a mean cost of $14,389 per surgery, surgical complications, nutritional deficiencies, and the need for drastic dietary changes [30].

In summary, although dietary weight loss can treat comorbidities associated with obesity, existing programs have shortcomings in sustainability, magnitude, adherence, and success rate. As such, a dietary weight loss program that can be used as a reliable treatment option is yet to be developed. Thus, we tested the feasibility of a new approach to dietary weight loss that does not rely on traditionally used methods. First, in place of common methods such as exclusion of food groups, strict weight-loss meal plans, and dieting products, iDip increases nutritional knowledge allowing participants to create their own weight loss diet by making informed and individualized food choices based on their preferences. Second, we monitored a target range of protein density per energy for participants to create a weight loss diet with increased protein while reducing the overall energy intake to meet an increased protein requirement during weight loss. Third, we replaced calorie counting/food journaling with daily weight monitoring and provision of a weekly progress chart to monitor energy balance.

## Materials and methods

This study was approved by the University of Illinois Institutional Review Board (#18069) on August 30^th^, 2017. The study was registered at the US National Institutes of Health (ClinicalTrial.gov) #NCT04605653. Registration was delayed due to a lack of knowledge on required clinical trial registration. The authors confirm that all ongoing and related trials for this intervention are registered. Participant recruitment started on September 1^st^, 2017, and the follow-up period ended on July 28^th^, 2018. Informed consent was obtained from all participants included in the study.

### Participant recruitment

Adults aged 18 to 64 years old and with a BMI >28 kg/m^2^ were recruited via posters, flyers, and word of mouth in Urbana-Champaign, Illinois, USA, and surrounding regions from September 2017 through January 2018. A power calculation was performed using an average standard deviation 7.0 for percentage weight loss at 12 months from similar dietary weight loss studies [31]. We set 5% weight loss as a primary outcome to be detected. The minimum sample number required with 95% confidence interval was 7.5. With n=10, we will be able to detect 5% difference when actual SD is 8.0. Alternatively, we will be able to detect 4.3% difference if SD is 7.0 as estimated. Based on a mean attrition rate of 22.1% from 60 studies [32], we recruited 14 participants assuming 30% attrition rate. A flow diagram of participant enrollment is shown in Fig 1.

**Fig 1.**
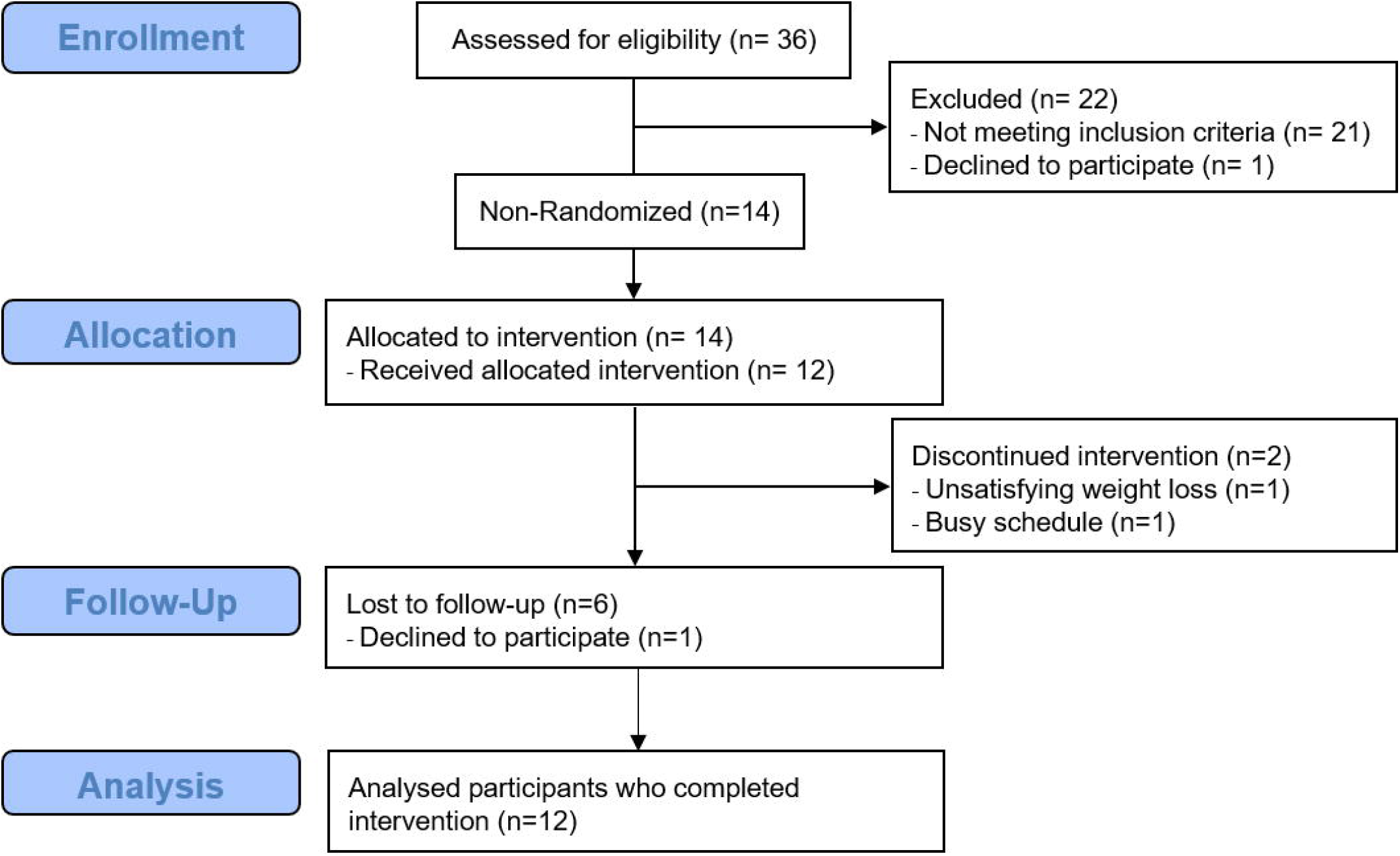
Flow diagram of the participants.

Inclusion criteria in addition to age and BMI were access to Wi-Fi at home, working email and smartphone, fluent English communicator, and willingness to self-weigh daily with a provided Wi-Fi scale for a minimum of 18 months. Exclusion criteria were the following: adults with self-reported severe metabolic, cardiovascular or musculoskeletal disease, insulin use, pregnancy, planned pregnancy, or lactating; or unwillingness to attend any of the educational sessions. Among eligible applicants, invitation was prioritized to best reflect the gender and racial composition of the community. Selected applicants were invited for an individual meeting for an additional screening process at the University of Illinois at Urbana-Champaign (UIUC). After receiving explanations about the study, a Food Frequency Questionnaire (FFQ) was administered, and anthropometric measurements were collected. Informed consent was obtained, and a Wi-Fi scale was provided. Other than a Wi-Fi scale, no monetary compensation was provided to the participants for participating in the study.

### Study design

The iDip study utilized a non-randomized, single-arm, before-and-after study design with a cohort of participants and comprised of 12 months of intervention followed by six months of follow-up. The objective was to test the feasibility of the informed decision-making approach to dietary weight loss by utilizing two forms of quantitative visual feedback for dietary improvement and weight loss progress. The iDip consisted of four weekly sessions (month one), 12 biweekly sessions (months two-six), and six monthly sessions (months seven-12). Nineteen 60-minute group-based diet improvement sessions and three 30-minute individual advising sessions were delivered by graduate students with registered dietitian credentials at UIUC from January 2018 to January 2019. All group sessions were focused on building knowledge and skills to make informed food selections. Weekly sessions in month one focused on the basic strategy of weight loss; bi-weekly sessions in months two-six focused on protein, fiber, physical activity, weight loss plateauing, and weight maintenance; and months seven-12 of monthly sessions focused on healthy eating and micronutrients beyond weight management (S1 Appendix). All participants received two types of feedback: 1) weekly weight chart and 2) protein-fiber (PF) plot (Fig 2) of their 24-hour diet record analysis at one, two, three, five, seven and 12 months and pre-and-post FFQs to assist individuals in monitoring their weight and dietary pattern, respectively. PF plot was the sole diet feedback and analysis that participants received. No food groups were excluded from participants’ diets, and even no dieting products nor recipes were distributed. Weight and dietary changes were evaluated during individual sessions, and participants were advised using our new approach.

**Fig 2.**
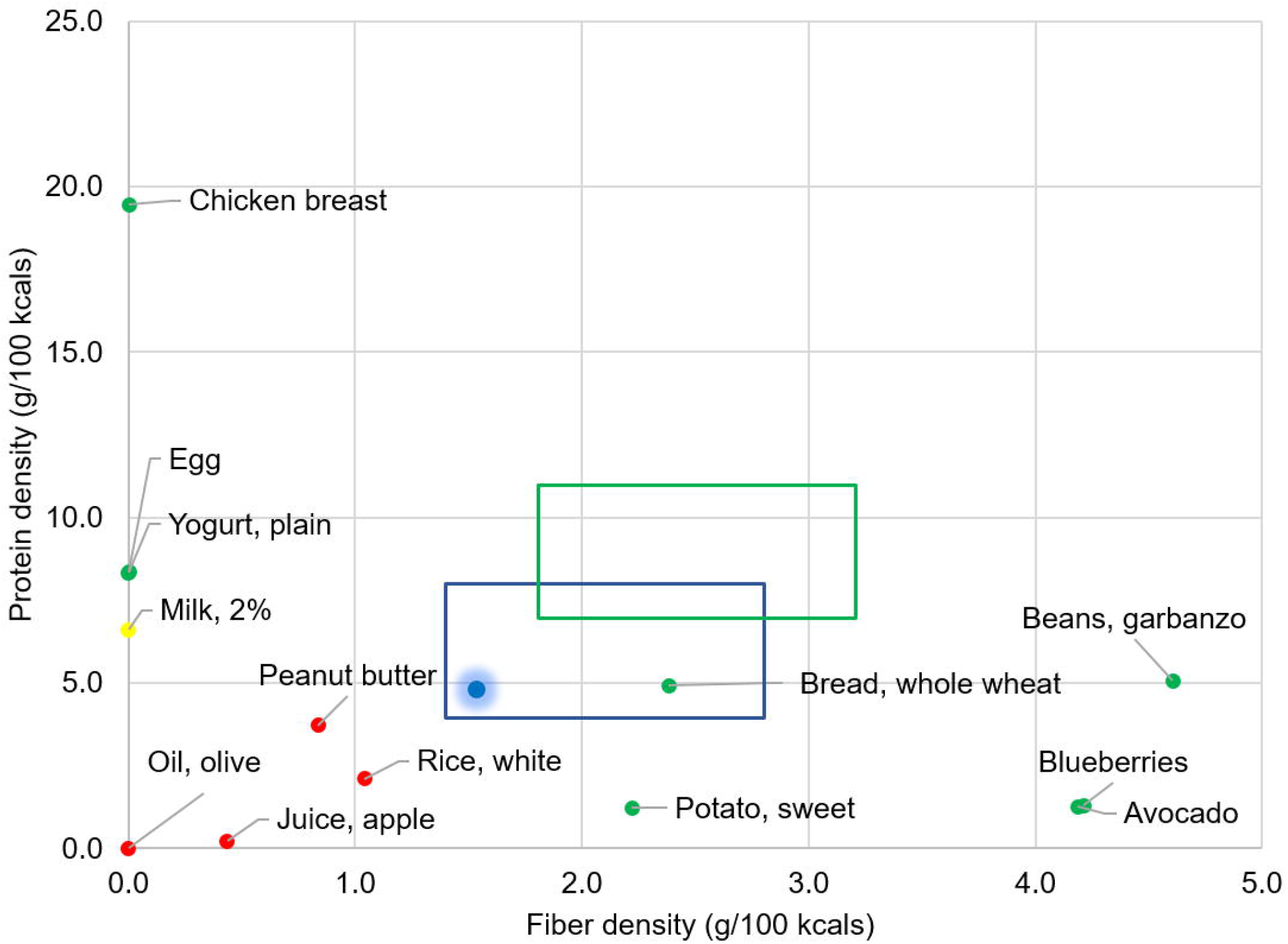
Protein and fiber plot with example food items. Foods with green dots are dense in protein or fiber and can be easily fit in the target box when combined, whereas foods with red dots cannot be due to low protein and fiber densities. A yellow food has protein and fiber density between green and red foods. A blue dot with a shadow shows where a meal falls when all foods are combined. Blue box, weight maintenance target; green box, weight loss target.

### Development of novel learning tools

#### A quantitative nutrient display method

The PF plot (Fig 2) is a two-dimensional data-visualization method for easy comparison of quantitative nutrition values in multiple foods designed to improve food selection based on protein and fiber density of foods [33]. The PF plot was developed by our lab, and its efficacy in aiding restaurant customers in selecting healthier menus has been demonstrated previously [33]. Protein and fiber are critical nutrients for weight management as adequate protein intake protects muscle mass from degradation during weight loss [22, 34], and consuming sufficient protein and fiber decreases excessive calorie consumption [35-37]. The fat to carbohydrate ratio was disregarded in this study as previous randomized-controlled trials have not shown that long-term weight loss favors a low-carbohydrate and high-fat diet or vice versa [38-40].

Protein and fiber targets of the PF plot are derived from the Acceptable Macronutrient Distribution Ranges (AMDR) and Adequate Intakes (AI) for weight maintenance in adults, respectively, as previously described [33]. Briefly, for protein, the AMDR is 10-36% daily intake; and for fiber, the AI is 14 g/1000 kcal daily [15]. For the weight maintenance box (blue): using 16-32% kcals from protein, we set 4-8 g/100 kcal as protein target; and for fiber, 1.4-2.8 g/100 kcal (Fig 2). The weight loss target (green) was set assuming a caloric reduction of minimum 25% of the daily requirement as weight loss requires a negative energy balance. By adjusting for 25% calorie reduction and 1.25x increase in protein requirement [22, 23], we set a protein range for weight loss of 7-11 g/100 kcal. Using 14 g fiber/1000 kcals and factoring 25% reduction of caloric intake, the resulting range was set as 1.8-3.2 g/100 kcals (Fig 2). The PF plot with a target box of protein and fiber density for weight loss was used as a key tool for food selection by participants throughout the study in both educational sessions and individual diet analysis.

### Energy balance monitoring

In the iDip, calorie counting and daily food journaling were replaced with daily self-weighing and weekly progress summary charts to monitor energy balance. As mentioned in the Introduction, calorie counting and food log methods are conventional in weight management, but these methods are time-consuming and prone to error, resulting in high attrition rates [19-21]. Daily self-weighing using a Wi-Fi-enabled scale (Weight Gurus, Greater Goods LLC, MO) provided a reliable energy balance estimation that requires little time. Participants were provided with a weekly weight feedback chart showing weight, a six month goal weight and an intended weight loss rate in the format shown in Fig 3.

**Fig 3.**
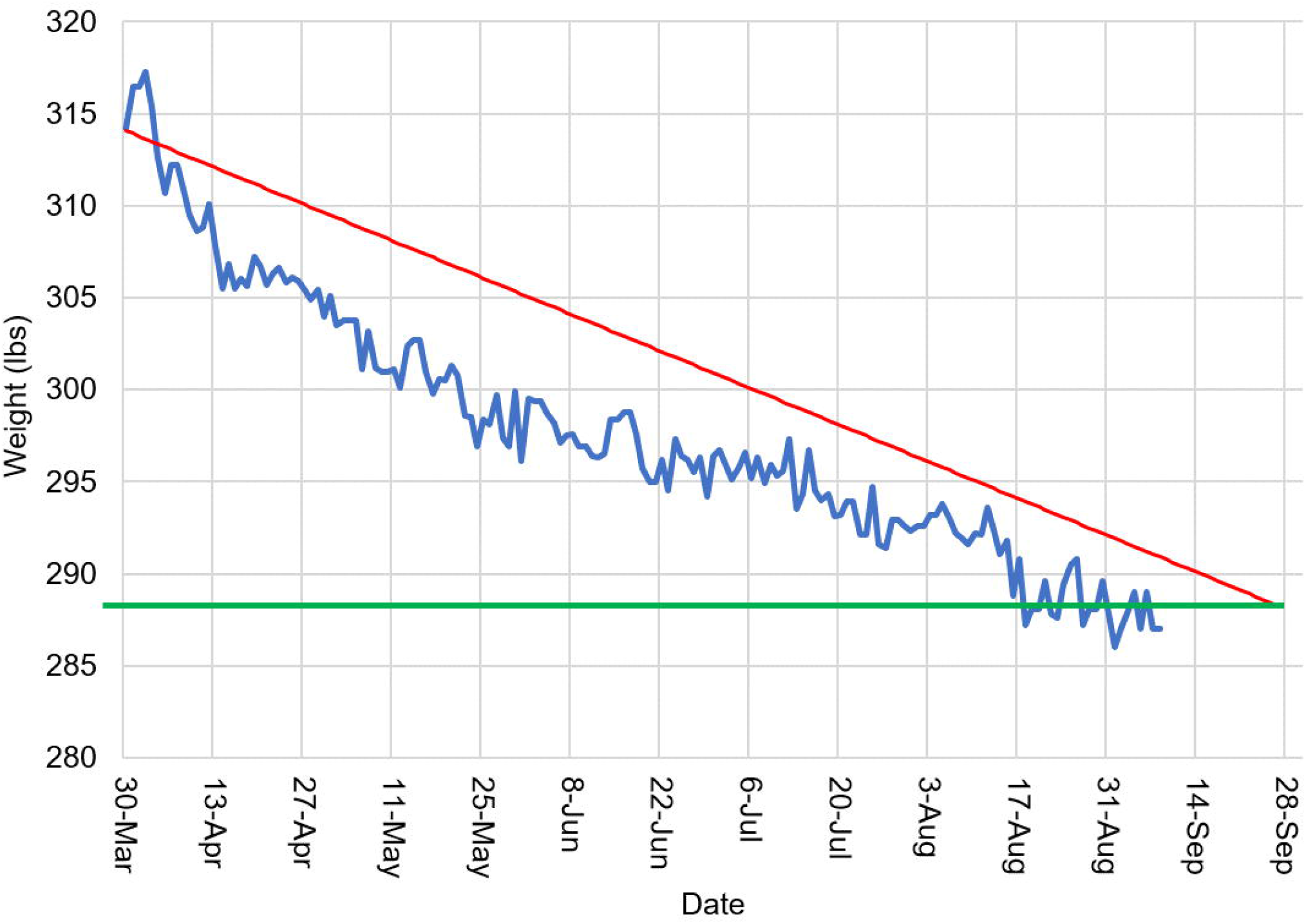
A weekly weight chart feedback. The red line represents the target weight loss slope (1 lb. loss (0.45 kg) per week) over time, the blue line represents actual daily weights, and the green line represents the target of 25 lbs. (11.3 kg) weight loss (based on a six month weight loss goal of 1 lb. (0.45 kg) loss per week).

### Outcome measures

#### Anthropometrics measures

Anthropometrics measurements were the primary outcomes, and entailed height, weight, waist, and hip circumferences. These were obtained at baseline and at 12 months. Height was recorded to the nearest 0.25 inches with shoes removed via stadiometer (Seca 700, Hanover, MD), and weight was measured to the nearest 0.1 lbs in light clothing on the same instrument. BMI was calculated using measured height and weight. In addition, weights were monitored and obtained daily via a Wi-Fi scale. Waist and hip circumferences were measured using a Retractable Tape Measure (Gulik II, Gay Mills, WI) and recorded to the nearest 0.1 cm. Waist to hip ratio was obtained based on measured waist and hip circumferences.

#### Dietary assessment

Dietary changes were the secondary outcomes assessed in this study. A paper version of the validated EPIC-Norfolk FFQ questionnaire (S2 Appendix) was used at baseline and at 12 months to assess dietary patterns [41]. Twenty-four-hour dietary records were administered at one, two, three, five, seven and 12 months to evaluate dietary changes. The results were returned to the participants in the form of a PF plot. The US Department of Agriculture National Nutrient Database for Standard Reference (https://fdc.nal.usda.gov/) and manufacturer information were used to calculate protein and fiber density.

#### Survey measures

An exit survey was conducted at the end of the 12 month intervention. A questionnaire containing 14 items was developed to explore personal gain, program content, program materials and feedback, engagement, and overall evaluation of the program. Possible responses for each question were “Definitely Yes,” “Yes,” “A little bit,” “Neutral,” “Not really,” “No,” “Definitely Not.” A score was provided for each response from one to seven, with seven being “Definitely Yes”.

### Data analysis

Descriptive analysis was used to summarize demographic characteristics. Weight loss of ≥5% from the baseline was considered meaningful as it has been shown to reduce risks associated with obesity [42]. Paired t-test analysis determined the pre- and post-intervention differences in outcome measures for the program completers. Pearson correlation analyses were performed to determine associations between body weight, protein density, and fiber density. Cohen’s d was computed by single sample z-test. Normality of data was tested using Shapiro-Wilk. Repeated measure ANOVA was used to compare 24-hour dietary records. All statistical analyses were performed using Microsoft Office Excel 2016 and R Computing (Version 3.6.1 © 2019), and p<0.05 was considered statistically significant.

## Results

### Participant characteristics

Fourteen participants started the study (Fig 1). The mean age was 44.6±3.0 years, and mean BMI was 36.7±1.7 kg/m^2^. The demographics and characteristics of participants are presented in Table 1. Enrolled participants had medical histories of hypertension (36%), hypercholesterolemia (21%), thyroid issues (21%), skeletal problems (14%), sleep apnea (14%), and hyperlipidemia (7%). Participants had prior dieting experiences including Weight Watchers (71%), Jenny Craig (14%), liquid diets (14%), Ideal Protein (14%), low carbohydrate (14%), South Beach (7%), low fat (7%), low calories (7%), cabbage soup diet (7%), Atkins (7%), 21-day fix (7%), Human Chorionic Gonadotropin (HCG) diet (7%), and Transformation diet (7%). No significant differences were found in any demographic data between top and bottom groups in weight loss.

**Table 1.** Demographic data of participants.

|  | **Enrollees (%)**<br>N=14 | **Completers (%)**<br>N=12 |
| --- | --- | --- |
| **Age in years** |  |  |
| 18-34 | 14.3 | 8.3 |
| 35-50 | 50.0 | 50.0 |
| 51-65 | 35.7 | 41.7 |
| **Gender** |  |  |
| Male | 21.4 | 25.0 |
| Female | 78.6 | 75.0 |
| **Marital Status** |  |  |
| Married | 64.3 | 75.0 |
| Single | 35.7 | 25.0 |
| **Ethnicity** |  |  |
| White | 57.1 | 50.0 |
| African American | 42.9 | 50.0 |
| **Educational level** |  |  |
| Graduate degree | 50.0 | 58.3 |
| Bachelor's degree | 42.9 | 41.7 |
| High school | 7.1 | 0.0 |

### Retention

Fourteen participants started the program. Participants who were unable to attend the regular sessions received make-up sessions. One participant dropped out after four weeks, and the other participant dropped out after eight weeks, yielding 85.7% retention. The reasons for leaving the study were as follows: too busy to focus on weight loss (n=1) and unsatisfying weight loss (n=1). During the six-month follow-up period, six out of 12 participants continued to weigh daily.

### Change in anthropometric measures

Anthropometric changes following the intervention are presented in Table 2 and Figures 4 and 5. At 12 months, six participants (50%) successfully lost >5% of their initial body weights (Fig 4). Of the unsuccessful participants (n=6), four participants did not reach 5% weight loss during the 12 months, whereas two participants regained lost weight after six months (Fig 4). On average, participants (n=12) achieved a significant weight loss of -5.4%±1.7% at 12 months (p <0.01) from the baseline (Fig 5). Cohen’s d for 12 month weight loss was -0.21 indicating a small, negative change in weight. A large difference in weight loss outcome was observed among participants at 12 months (p<0.001); the top half (n=6) showed a significant loss from the baseline reaching - 9.7±1.7%, whereas the bottom half (n=6) lost -1.0±1.4% (Fig 5). The difference between the two groups widened as time progressed from the start to 12 months, particularly during the last five months (Fig 5).

**Table 2.**
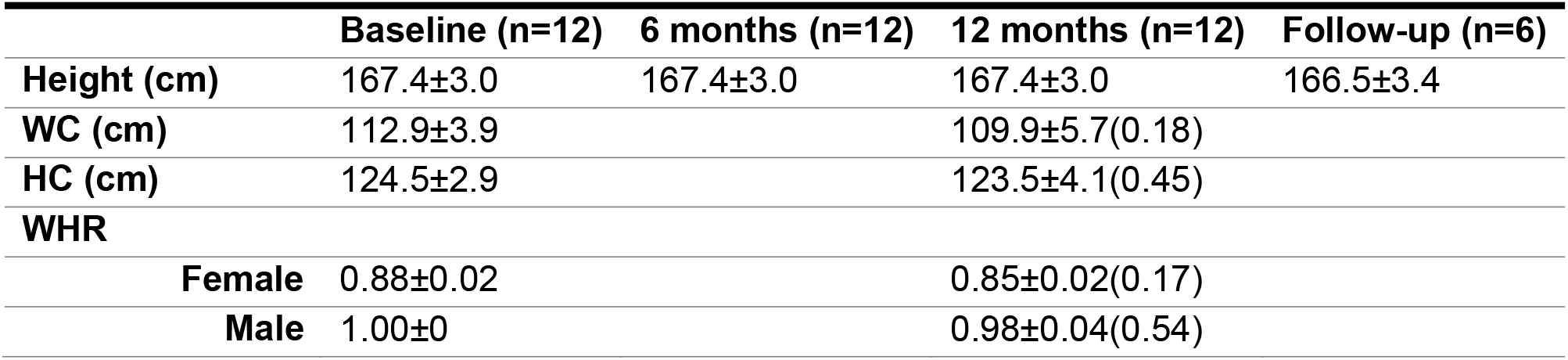
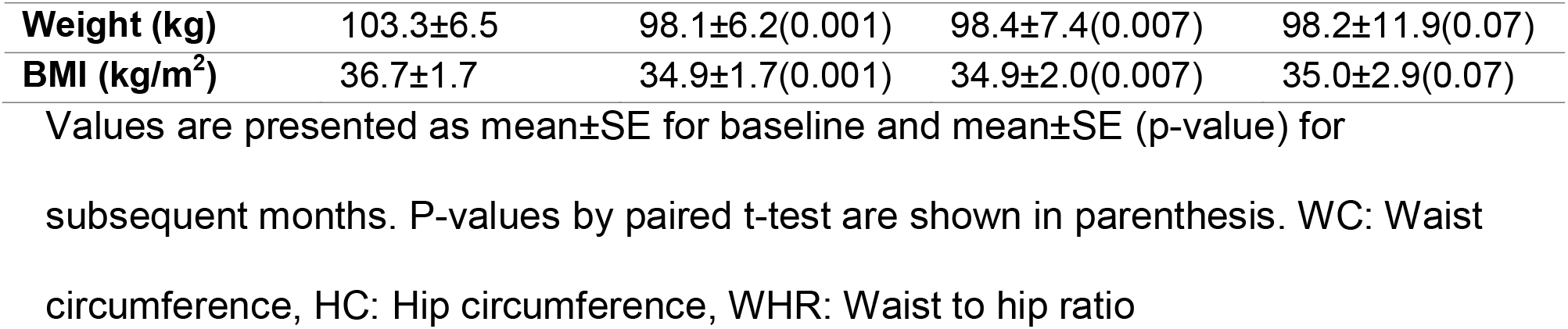
Characteristics of participants who completed dietary improvement sessions.

**Fig 4.**
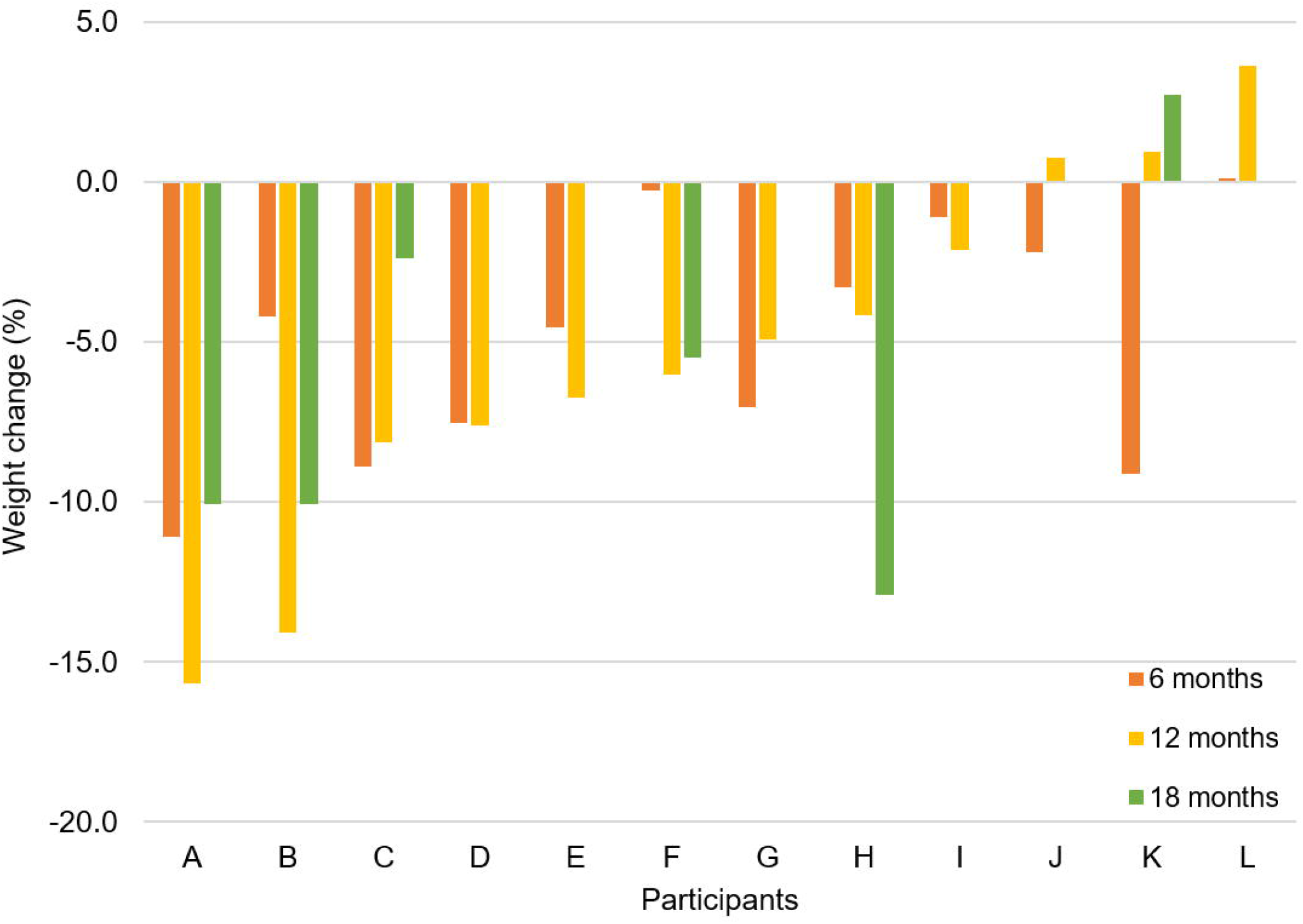
Weight changes (%) of each participant at six, 12, and 18 months. The data are ordered by magnitudes of weight loss at 12 months.

**Fig 5.**
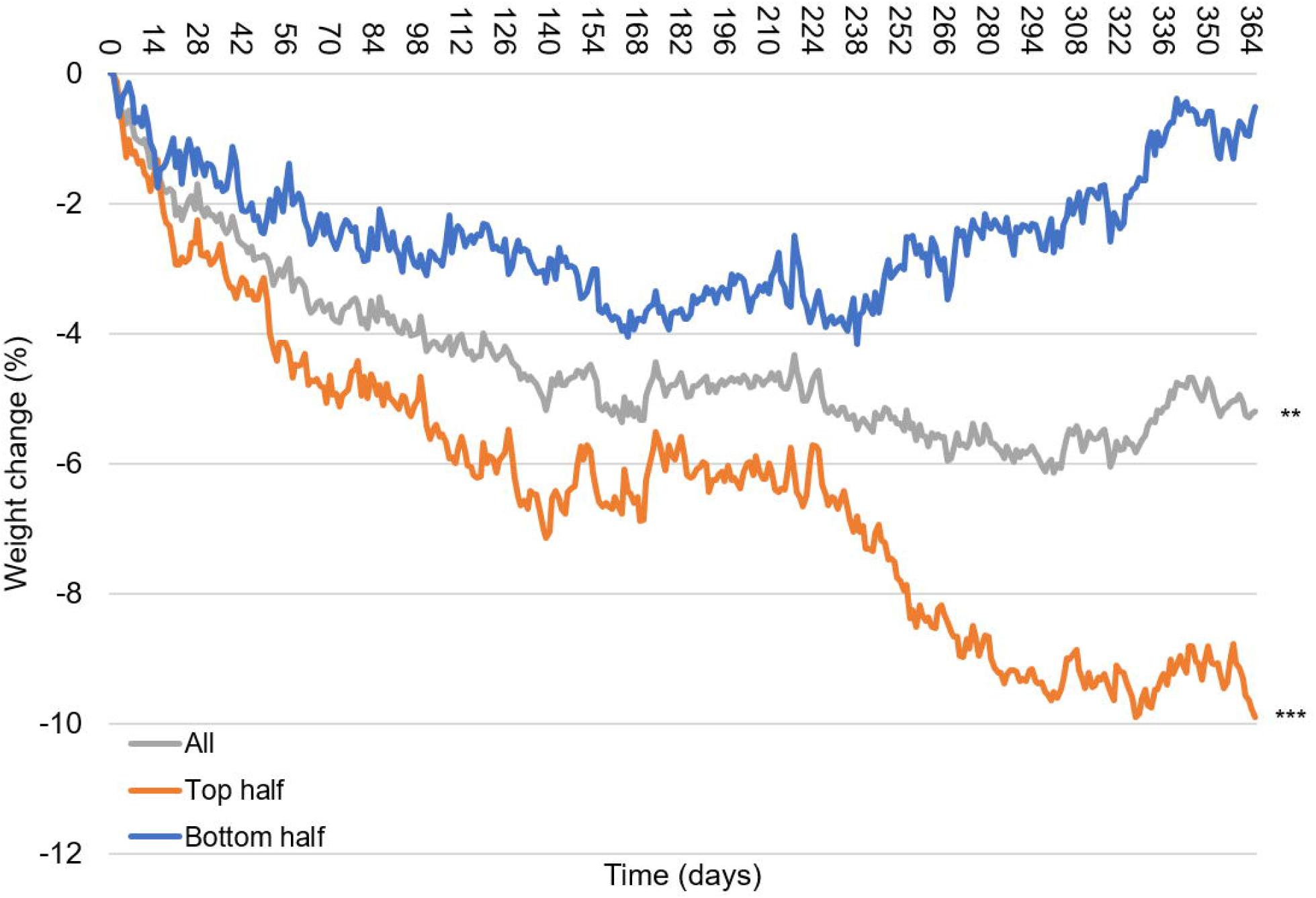
Weight changes (%) of all, top-half and bottom-half groups over 12 months. Participants were divided into two groups of equal size (n=6 each) based on the weight changes from baseline at 12 months. **p<0.01, ***p<0.001 at 12 months compared with baseline.

### Change in dietary components

Changes in diets over 12 months based on FFQ and 24-hour dietary records are shown in Fig 6. A significant increase in fiber density (p=0.02) but not protein density was found from the FFQ. Twenty-four-hour records showed a significant improvement in protein density in all 5 subsequent records when compared with the first month record, whereas fiber density increased only at three months (p=0.02). Next, 24-hour records were compared between the groups of the bottom and top half in weight loss at 12 months (Fig 7). While only one out of six records of the bottom half was inside the weight maintenance box, four out of six records of the top half stayed within the weight maintenance box. A significant difference in fiber density was observed between the top half and bottom half (p=0.04) although there was no difference in protein density (p=0.19) (Fig 7). There was no significant difference in protein density between the two groups at any time point.

**Fig 6.**
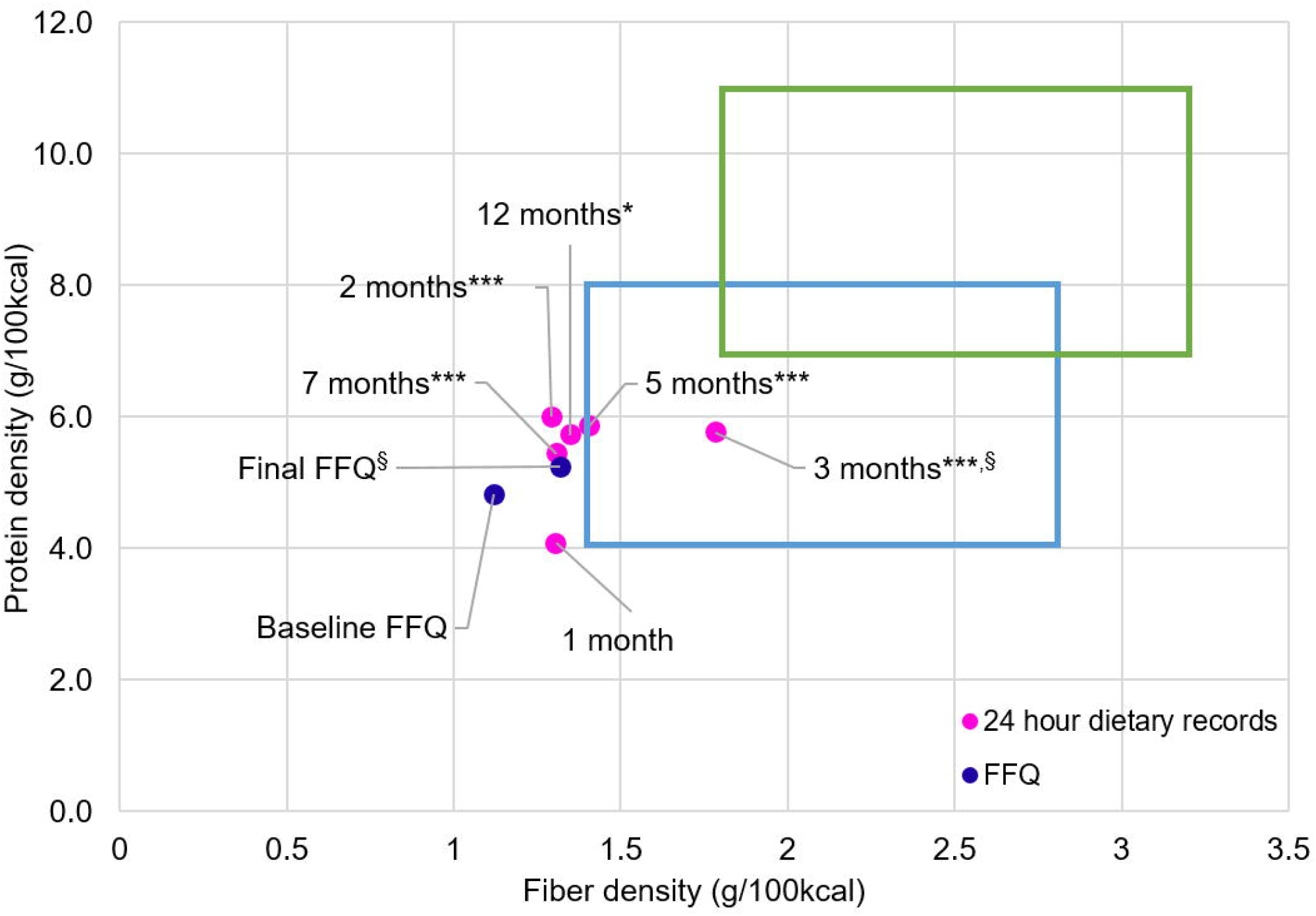
Changes in protein and fiber density of the whole group assessed by FFQ and 24-hour records. Data are presented as mean. FFQ were collected at the baseline (n=12) and the end of the trial (n=11). Twenty-four-hour records were collected at one – seven months (n=12) and at 12 months (n=10). Changes in protein density and fiber density were compared between baseline and 12 months in FFQ, and between the first month and two-12 months in 24-hour records. *p=0.02, ***p<0.001 in protein density; §p=0.02 in fiber density by paired t-test.

**Fig 7.**
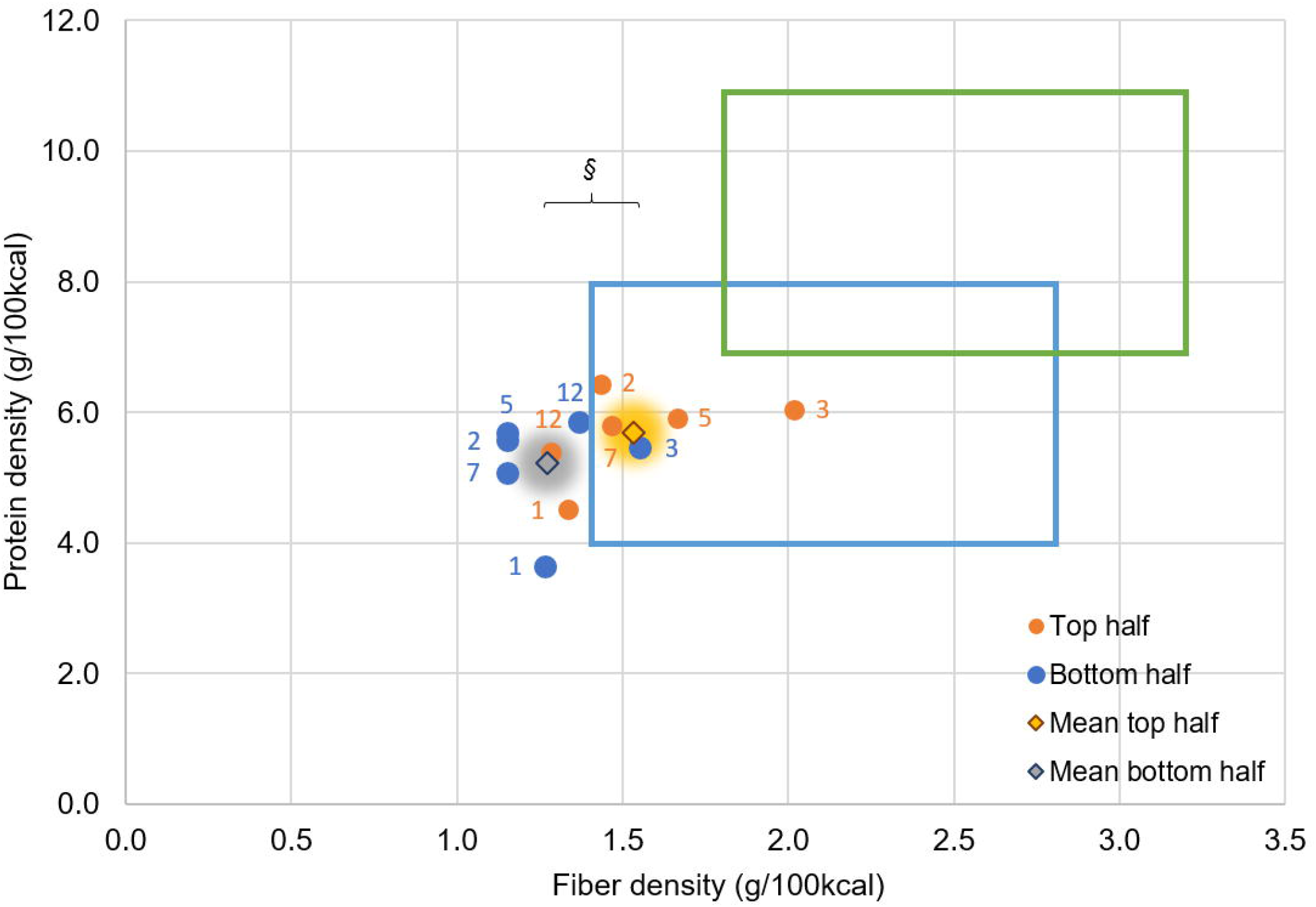
Changes in protein and fiber density of top and bottom half groups in weight loss at 12 months. Twenty-four-hour records conducted at one, two, three, five, seven and 12 months. Dietary records were divided to two groups based on top half (n=6) and bottom half (n=6) of weight loss at 12 months. Data are presented as mean. §p=0.04 significant difference in fiber density between two groups by repeated measure ANOVA when all time points of 24-hour records were combined (n=35 for each group).

### Correlation among weight loss and protein and fiber density

Correlations between diet and weight changes were tested to identify possible determinants of weight loss success (Fig 8). A significant correlation was found between weight loss at three months and weight loss at 12 months, suggesting participants who lost weight during the initial three months continued to lose more (p=0.03, r_p_=0.64). A positive correlation was observed between protein and fiber densities at three months (p=0.02, r_p_=0.64). A positive correlation between protein density at three months and its density at six months was also observed (p=0.002, r_p_=0.80). No significant associations were found between weight loss and either protein or fiber density at any months tested.

**Fig 8.**
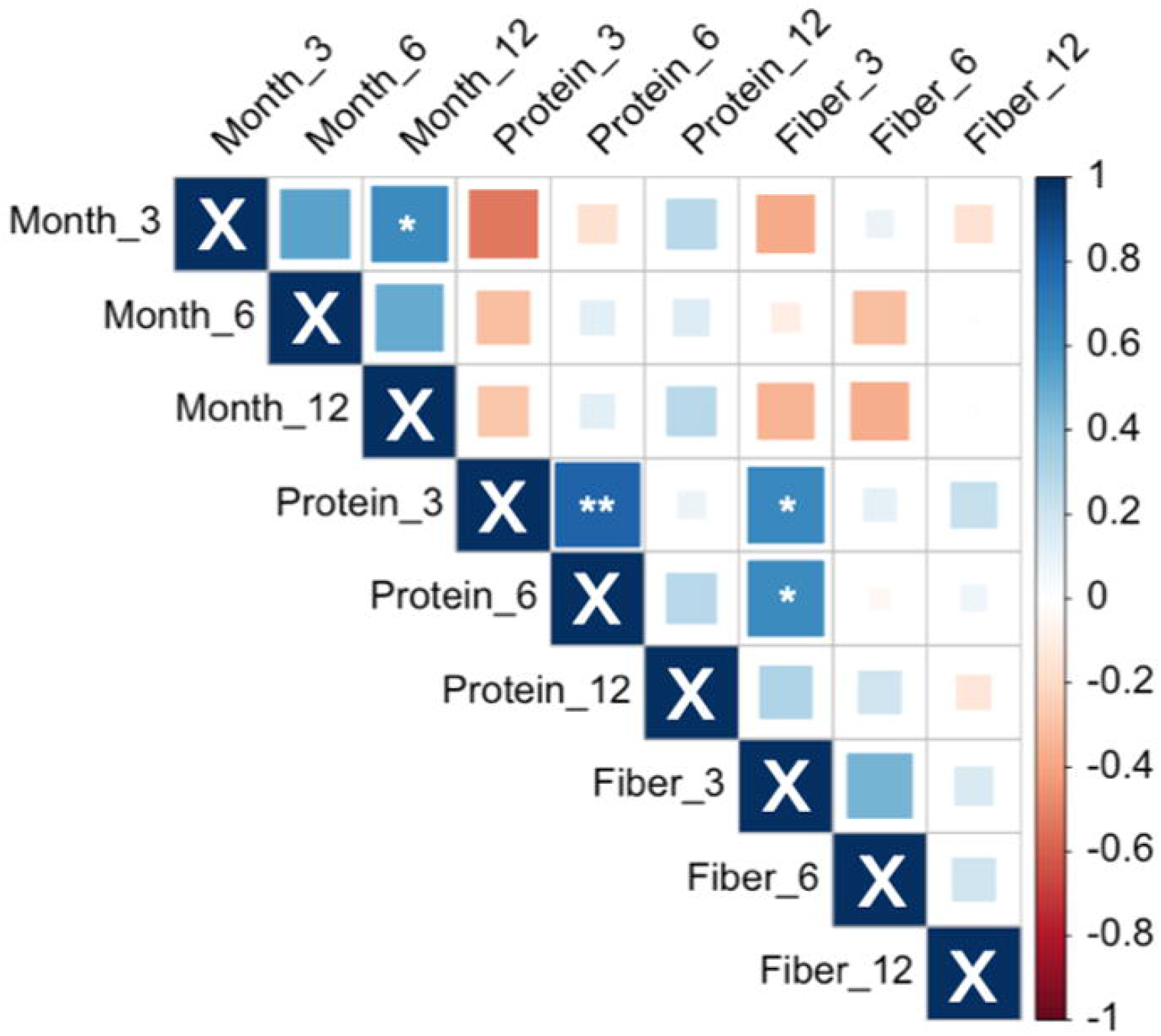
Correlation matrix showing the interrelationship among weight loss and protein and fiber density at three months, six months, and 12 months. Positive correlations are displayed in blue and negative correlations in red. *p<0.05, **p<0.01.

### Exit survey

The results of the exit survey at 12 months are shown in S3 Appendix. Questions one and three suggested that participants who completed the program found it beneficial for applying skills such as interpreting nutrition information to maintain healthier eating behaviors. A high score was received in program content with instructors being knowledgeable (question 5) and available to offer support (question 8). Finally, participants reported that the visual feedbacks, including the PF plot and weekly weight chart, were easy to understand and helpful (questions 9 and 10). The overall findings from the survey suggested that iDip equipped participants with valuable nutrition knowledge and skills, which enabled some individuals to adhere to dietary changes. However, the scores of questions 2 and 11 showed that the program was lacking in some aspects of satisfaction in overall engagement and individual goal achievements.

## Discussion

This non-randomized and single-arm trial demonstrated the feasibility of a new approach to weight loss that utilized two key visual feedback systems: the PF plot and the weekly weight chart. These quantitative feedbacks allowed participants to self-select dietary modifications and monitor their energy balance without calorie counting. iDip uniquely used no calorie counting, meal plans or recipes, meal replacements, or food exclusions. At 12 months, mean weight loss was -5.4±1.7% (p <0.01), and 50% of participants achieved >5% weight loss.

### Innovations

iDip was aimed to increase dietary flexibility and easily monitor energy balance, two shortcomings of many existing weight loss programs. To increase dietary flexibility, iDip provided nutrition education and individualized dietary feedback in the form of a PF plot to empower participants to develop a weight loss diet suitable to their taste preferences and lifestyle. Participants were able to create and advance a weight loss diet based on the PF plot. Participants were not given specific meal plans, meal replacement products, or recipes; rather, all dietary modifications were selected and implemented by individuals with guidance from individual counseling and educational sessions delivered by dietitians. Importantly, no macronutrient or food group was excluded or limited in contrast to the practice commonly used in dieting programs, such as the Atkins diet. Also, iDip did not place any focus on the fat and carbohydrate ratios in a diet as modifying fat and carbohydrate ratios has been reported to be ineffective for long-term weight loss [10, 13]. Participants were encouraged to self-select to limit intake of foods that contain minimal fiber and protein, but they were not instructed to exclude any particular foods from their diet. Thus, all diets and macronutrient distributions differed by participants.

The iDip study participants significantly lost weight by creating their weight-loss diet based on feedback in the form of a PF plot. Previously in a 12-week university cafeteria study, we tested if providing nutrient information in a PF plot format could assist customers in making an informed, healthier choice [33]. In the study, no nutritional information was provided during weeks one-three and seven-nine, and a Nutrition Facts Panel and a PF plot were posted on identical menu items, during weeks four-six and weeks 10-12, respectively. Sales data, the primary outcome, showed that when the PF plot was displayed, the protein content and protein + fiber content of purchased meals was significantly higher than no posting periods, whereas the Nutrition Facts Panel had no effect. These results have shown the PF plot’s ease of understanding and its ability to improve consumer food choices [33]. In the current study, all educational session materials and individual diet feedback were provided in the form of a PF plot, demonstrating the applicability of the PF plot to creating a weight-loss diet by individual participants.

iDip utilized daily self-weighing on a Wi-Fi-enabled scale to provide easy and reliable energy balance monitoring without calorie counting or daily food journals. Self-monitoring is a critical factor in behavior changes for weight loss [43, 44]. Bertz et al. showed daily self-weighing and weekly summary feedback can limit weight gain [45]. We provided a cumulative weight loss progress chart every week, eliminating the need for calorie counting entirely and providing means for self-monitoring. The purpose of the six 24-hour records administered was to evaluate progress in dietary improvement, not to monitor energy balance.

iDip had a retention rate of 85.7%, indicating likeability of our program and its flexible dietary approach. This high retention was achieved without monetary compensation. Participants were given the Wi-Fi scale (value ∼ $60) to keep after completing the study. The dietary flexibility, self-selection of foods, and no need for daily calorie counting may have at least in part contributed to the low attrition.

### Limitations

We chose the minimum number of participants to achieve the study objective of testing the feasibility of a novel dietary weight loss approach, not to test the robustness of the program as a clinical treatment option. Despite achieving a clinically significant mean percent weight loss of >5%, we observed some limitations. First, a success rate of 50% of participants achieving >5% weight loss at 12 months is not sufficient to be used as a reliable treatment program. Also, a greater magnitude of weight loss will be needed for iDip to become a clinical treatment option. Second, the follow-up period (six months) was too short to assess long-term sustainability. At 18 months, 50% of the cohort was lost to follow-up and were no longer weighing regularly, suggesting that half of the participants regarded daily weighing as a requirement of the program, not an essential part of weight management. Among the remaining participants, not all were successful in maintaining their weight between 12 and 18 months, indicating the dietary changes made during the program were not sustained in some. iDip aimed to improve long-term sustainability through non-exclusion of certain food groups and self-selected approach; thus, future studies will include a longer follow-up period with better implementation of sustainable dietary changes and daily weighing for monitoring energy balance. Third, body composition changes were not measured or tracked throughout the study. However, with the focus on increased protein density, we expect little muscle mass loss. Lastly, changes in health parameters, such as blood lipid and glucose levels and changes in medications associated with obesity and other comorbidities, were not tracked. Although a randomized and controlled trial is standard for clinical studies, a large-scale, randomized, controlled weight-loss trial has shown that the weight of the placebo group was unchanged during one year, whereas the metformin and lifestyle modification groups lost significant weight [46]. Therefore, this landmark study suggests that the lack of a no-treatment control group would not pose a major limitation in interpreting the weight loss outcome of our study.

### Future directions

This study showed proof of concept for the non-exclusion of certain food groups and self-selected approach to dietary weight loss. Based on this success, steps of future studies will include: 1) increase the magnitude of mean percent weight loss; 2) improve the overall success rate of participants achieving >5% weight loss; 3) monitor body composition to minimize loss of skeletal muscle mass; 4) calculate intervention cost; and 5) collaborate with other health professionals to track health markers, such as blood lipids, and address medical and psychological barriers to weight loss. In the long term, a large-scale, multi-center trial with a comparable control group is required to test general applicability and effectiveness of the program as a treatment option for obesity.

## Conclusions

In conclusion, this non-randomized and single-arm trial has shown the feasibility of the informed decision-making approach for dietary weight loss by utilizing two forms of quantitative visual feedback. One was the PF plot to monitor protein per energy and fiber per energy intake, and the other was the daily weight chart to monitor energy balance. Participants understood these visual weight management tools well. Participants significantly improved fiber density from the baseline FFQ to the final FFQ, and protein density significantly increased from the baseline 24-hour record at several time points in the study. The participants achieved a mean percent weight loss of - 5.4±1.7%, and half of them successfully lost ≥5% baseline weight. The dietary flexibility and self-selection of foods may have contributed to a low attrition rate. Future studies are warranted to improve success rate, magnitude of weight loss and long-term sustainability.

## Supporting information

Supplemental Checklist 1

Supplemental Checklist 2

Study Protocol

## Data Availability

Data are available without restriction.

https://doi.org/10.13012/B2IDB-4710255_V1

## Supporting information

**S1 Appendix. iDip educational session topics**

**S2 Appendix. Food Frequency Questionnaire**

**S3 Appendix. Exit survey outcomes**

**S1 Checklist. CONSORT for pilot and feasibility study**

**S2 Checklist. TIDieR Checklist**

**S1 Protocol. Clinical trial protocol**

