## Supplemental Appendix 1 for "A feasibility study to test a novel approach to dietary weight loss with a focus on assisting informed decision making in food selection"

**S1 Table. iDip educational session topics**

| Dietary Improvement Session Topics | |
|---|---|
| **1** | FFQ, starting weight loss |
| **2** | Weight monitoring |
| **3** | 24-hour dietary record 1 |
| **4** | Individual advising 1 |
| **5** | Protein 1 |
| **6** | Establishing routine and 24 hour dietary record 2 |
| **7** | Fiber 1 |
| **8** | Individual advising 2 |
| **9** | Physical activity |
| **10** | Protein 2 |
| **11** | Fiber 2 |
| **12** | Trouble shooting for slowing down |
| **13** | Peer experience sharing |
| **14** | Weight maintenance: Difference between weight loss and maintenance |
| **15** | Weight maintenance: Building a Healthy Plate |
| **16** | Weight maintenance: Hidden Kcal Contributors and Diet Foods |
| **17** | Barriers to Healthy Eating |
| **18** | Fats |
| **19** | Individual advising 3 |
| **20** | Salt and Potassium |
| **21** | Eat the Rainbow: The Importance of Vitamins and Minerals |
| **22** | Final review |
