## Supplemental Appendix 2 for "A feasibility study to test a novel approach to dietary weight loss with a focus on assisting informed decision making in food selection"

### FOOD FREQUENCY QUESTIONNAIRE

**This questionnaire asks for some background information about you, especially about what you eat.**

**Please answer every question. If you are uncertain about how to answer a question then do the best you can, but please do not leave a question blank.**

1. **YOUR DIET LAST YEAR**

For each food there is an amount shown, either a “medium serving” or a common household unit such as a slice or teaspoon. Please put a tick (✓) in the box to indicate how often, **on average**, you have eaten the specified amount of each food **during the past year**.

#### EXAMPLES:

For white bread the amount is one slice, so if you ate 4 or 5 slices a day, you should put a tick in the column headed “4-5 per day”.

| FOODS AND AMOUNTS | AVERAGE USE LAST YEAR | | | | | | | | |
|---|---|---|---|---|---|---|---|---|---|
| **BREAD AND SAVOURY BISCUITS** (one slice or biscuit) | Never or less than once/month | 1-3 per month | Once a week | 2-4 per week | 5-6 per week | Once a day | 2-3 per day | 4-5 per day | 6+ per day |
| White bread and rolls | | | | | | | | ✓ | |

For chips, the amount is a “medium serving”, so if you had a helping of chips twice a week you should put a tick in the column headed “2-4 per week”.

| FOODS AND AMOUNTS | AVERAGE USE LAST YEAR | | | | | | | | |
|---|---|---|---|---|---|---|---|---|---|
| **POTATOES, RICE AND PASTA** (medium serving) | Never or less than once/month | 1-3 per month | Once a week | 2-4 per week | 5-6 per week | Once a day | 2-3 per day | 4-5 per day | 6+ per day |
| Chips | | | | ✓ | | | | | |

For very seasonal fruits such as strawberries and raspberries you should estimate your average use when the fruits are in season, so if you ate strawberries or raspberries about once a week when they were in season you should put a tick in the column headed “once a week”

| FOODS AND AMOUNTS | AVERAGE USE LAST YEAR | | | | | | | | |
|---|---|---|---|---|---|---|---|---|---|
| **FRUIT** (1 fruit or medium serving) | Never or less than once/month | 1-3 per month | Once a week | 2-4 per week | 5-6 per week | Once a day | 2-3 per day | 4-5 per day | 6+ per day |
| Strawberries, raspberries, kiwi fruit | | | ✓ | | | | | | |

EXAMPLE

PLEASE PUT A TICK (✓ ) ON EVERY LINE

| MEAT AND FISH | Never or less than once a month | 1-3 per month | Once a week | 2-4 per week | 5-6 per week | Once a day | 2-3 per day | 4-5 per day | 7 per day |
|---|---|---|---|---|---|---|---|---|---|
| Beef: roast, steak, mince, stew or casserole (5oz.) | | | | | | | | | |
| Beef burgers (6oz.) | | | | | | | | | |
| Pork: roast, chops, stew or slices (5oz.) | | | | | | | | | |
| Lamb: roast, chops or stew (5oz.) | | | | | | | | | |
| Chicken or other poultry e.g. turkey (5oz.) | | | | | | | | | |
| Bacon (2oz. or 4 slices) | | | | | | | | | |
| Ham (3oz.) | | | | | | | | | |
| Corned beef, Spam, luncheon meats (4oz.) | | | | | | | | | |
| Sausages (5oz.) | | | | | | | | | |
| Liver (5oz.) | | | | | | | | | |
| Fried fish in batter, as in fish and chips (5oz.) | | | | | | | | | |
| Fish fingers, fish cakes (5oz.) | | | | | | | | | |
| Other white fish, fresh or frozen, e.g. cod, haddock, plaice, sole, halibut (5oz.) | | | | | | | | | |
| Oily fish, fresh or canned, e.g. mackerel, kippers, tuna, salmon, sardines, herring (5oz.) | | | | | | | | | |
| Shellfish, e.g. crab, prawns, mussels (5oz.) | | | | | | | | | |
| Fish roe, taramasalata (2oz.) | | | | | | | | | |
| **BREAD AND SAVOURY BISCUITS** | | | | | | | | | |
| White bread and rolls (2 slices) | | | | | | | | | |
| Brown bread and rolls (2 slices) | | | | | | | | | |
| Whole wheat bread and rolls (2 slices) | | | | | | | | | |
| Cream crackers, cheese biscuits (6 crackers) | | | | | | | | | |
| Crispbread, e.g. Ryvita (4oz.) | | | | | | | | | |
| **CEREALS** | | | | | | | | | |
| Oatmeal (1/2 cup) | | | | | | | | | |
| Breakfast cereal such as cornflakes, muesli etc. (1 cup) | | | | | | | | | |

PLEASE PUT A TICK (✓ ) ON EVERY LINE

| **POTATOES, RICE AND PASTA** | **Never or less than once a month** | **1-3 per month** | **Once a week** | **2-4 per week** | **5-6 per week** | **Once a day** | **2-3 per day** | **4-5 per day** | **7 per day** |
|---|---|---|---|---|---|---|---|---|---|
| Boiled, mashed potatoes (3.5oz.) | | | | | | | | | |
| French Fries (4oz.) | | | | | | | | | |
| Roast potatoes (3.5oz.) | | | | | | | | | |
| Potato salad (3.5oz.) | | | | | | | | | |
| White rice (1/2 cup) | | | | | | | | | |
| Brown rice (1/2 cup) | | | | | | | | | |
| White or green pasta, e.g. spaghetti, macaroni, noodles (1 cup) | | | | | | | | | |
| Whole wheat pasta (1 cup) | | | | | | | | | |
| Lasagna, moussaka (1 square) | | | | | | | | | |
| Pizza (1 large slice) | | | | | | | | | |
| **DAIRY PRODUCTS AND FATS** | | | | | | | | | |
| Sour cream (tablespoon) | | | | | | | | | |
| Double or Clotted cream (tablespoon) | | | | | | | | | |
| Low fat Greek yogurt (125g/ 4.4oz. carton) | | | | | | | | | |
| Full fat Greek yogurt (125g/ 4.4 oz. carton) | | | | | | | | | |
| Low fat yogurt, e.g. Vanilla (1/2 cup) | | | | | | | | | |
| Full fat yogurt, e.g. mixed berries (1/2 cup) | | | | | | | | | |
| Low fat, skim or non-fat milk (8oz.) | | | | | | | | | |
| Whole milk (8oz.) | | | | | | | | | |
| Cheese, e.g. Cheddar, Brie, Edam (medium serving/ 1.3oz.) | | | | | | | | | |
| Cottage cheese (2.4oz.) | | | | | | | | | |
| Eggs as boiled, scrambled, etc. (one) | | | | | | | | | |
| Quiche (3.5oz.) | | | | | | | | | |
| Low fat salad cream (tablespoon) | | | | | | | | | |
| Mayonnaise, salad cream (tablespoon) | | | | | | | | | |
| French dressing (tablespoon) | | | | | | | | | |

PLEASE PUT A TICK (✓ ) ON EVERY LINE

| **The following on bread or vegetables** | **Never or less than once a month** | **1-3 per month** | **Once a week** | **2-4 per week** | **5-6 per week** | **Once a day** | **2-3 per day** | **4-5 per day** | **7 per day** |
|---|---|---|---|---|---|---|---|---|---|
| Butter (teaspoon) | | | | | | | | | |
| Block margarine (teaspoon) | | | | | | | | | |
| Polyunsaturated margarine, e.g. Flora, sunflower (teaspoon) | | | | | | | | | |
| Low fat spread (teaspoon) | | | | | | | | | |
| **SWEETS AND SNACKS** | | | | | | | | | |
| Sweet crackers, e.g. Wafer (3 crackers) | | | | | | | | | |
| Cookies, e.g. plain, chocolate, ginger (one) | | | | | | | | | |
| Cake home baked, ready made (one slice/ 1.5oz.) | | | | | | | | | |
| Donut home baked, ready made (1.5oz.) | | | | | | | | | |
| Fruit pies, tarts, crumbles, home made, ready made (one slice/ 1.8oz.) | | | | | | | | | |
| Chocolate pudding (one container) | | | | | | | | | |
| Milk pudding (3.5oz.) | | | | | | | | | |
| Ice cream (125g carton/ 4.4oz.) | | | | | | | | | |
| Chocolates, single or square, e.g. Hershey's Milk Chocolate (4 squares) | | | | | | | | | |
| Chocolate snack bars, e.g. Snickers (one bar) | | | | | | | | | |
| Candy, sweets, toffees, mints (one pop) | | | | | | | | | |
| Sugar added to tea, coffee, cereal (teaspoon) | | | | | | | | | |
| Potato chips (one snack size bag) | | | | | | | | | |
| Peanuts or other nuts (1oz.) | | | | | | | | | |
| **SOUPS, SAUCES, AND SPREADS** | | | | | | | | | |
| Vegetable soups (bowl) | | | | | | | | | |
| Beef soups (bowl) | | | | | | | | | |
| Gravy (tablespoon) | | | | | | | | | |
| Tomato ketchup (tablespoon) | | | | | | | | | |
| Pickles (tablespoon) | | | | | | | | | |

PLEASE PUT A TICK (✓ ) ON EVERY LINE

| SOUPS, SAUCES, AND SPREADS | Never or less than once a month | 1-3 per month | Once a week | 2-4 per week | 5-6 per week | Once a day | 2-3 per day | 4-5 per day | 7 per day |
|---|---|---|---|---|---|---|---|---|---|
| Jam, marmalade, honey (teaspoon) | | | | | | | | | |
| Peanut butter (teaspoon) | | | | | | | | | |
| **DRINKS** | | | | | | | | | |
| Tea (cup) | | | | | | | | | |
| Coffee, instant or ground (cup) | | | | | | | | | |
| Coffee, decaffeinated (cup) | | | | | | | | | |
| Coffee creamer, e.g. Coffee-mate (teaspoon) | | | | | | | | | |
| Cocoa, hot chocolate (cup) | | | | | | | | | |
| Wine (glass) | | | | | | | | | |
| Beer (half pint) | | | | | | | | | |
| Port, sherry, vermouth, liquors (shot glass) | | | | | | | | | |
| Spirits, e.g. gin, brandy, whiskey, vodka (shot glass) | | | | | | | | | |
| Low or diet fizzy soft drinks (can) | | | | | | | | | |
| Fizzy soft drinks, e.g. Coca cola, lemonade (glass) | | | | | | | | | |
| Pure fruit juice (100%) e.g. orange, apple juice (glass) | | | | | | | | | |
| Fruit juice from concentrate (one can) | | | | | | | | | |
| **FRUIT** | | | | | | | | | |
| Apples (1 fruit) | | | | | | | | | |
| Pears (1 fruit) | | | | | | | | | |
| Oranges, satsumas, mandarins (1 fruit) | | | | | | | | | |
| Grapefruit (half) | | | | | | | | | |
| Bananas (1 fruit) | | | | | | | | | |
| Grapes (medium serving) | | | | | | | | | |
| Melon (1 slice) | | | | | | | | | |
| Peaches, plums, apricots (1 fruit) | | | | | | | | | |
| Strawberries, raspberries, kiwi fruit (medium serving) | | | | | | | | | |
| Canned fruit (medium serving) | | | | | | | | | |
| Dried fruit, e.g. raisins, prunes (1/4 cup) | | | | | | | | | |

**PLEASE PUT A TICK (✓ ) ON EVERY LINE**

| VEGETABLES | Never or less than once a month | 1-3 per month | Once a week | 2-4 per week | 5-6 per week | Once a day | 2-3 per day | 4-5 per day | 7 per day |
|---|---|---|---|---|---|---|---|---|---|
| Carrots (1.8oz.) | | | | | | | | | |
| Spinach (1.8oz.) | | | | | | | | | |
| Broccoli, spring greens, kale (1.8oz.) | | | | | | | | | |
| Brussels sprouts (1.8oz.) | | | | | | | | | |
| Cabbage (1.8oz.) | | | | | | | | | |
| Peas (1.8oz.) | | | | | | | | | |
| Green beans, broad beans, runner beans (1.8oz.) | | | | | | | | | |
| Zucchini (1.8oz.) | | | | | | | | | |
| Cauliflower (1.8oz.) | | | | | | | | | |
| Parsnips, turnips, swedes (1.8oz.) | | | | | | | | | |
| Leeks (1.8oz.) | | | | | | | | | |
| Onions (1.8oz.) | | | | | | | | | |
| Garlic (teaspoon) | | | | | | | | | |
| Mushrooms (1.8oz.) | | | | | | | | | |
| Sweet pepper, e.g. red, yellow (1.8oz.) | | | | | | | | | |
| Bean sprouts (1.8oz.) | | | | | | | | | |
| Green salad, lettuce, cucumber, celery (1.8oz.) | | | | | | | | | |
| Watercress (1.8oz.) | | | | | | | | | |
| Tomatoes (1.8oz.) | | | | | | | | | |
| Sweet corn (1.8oz.) | | | | | | | | | |
| Beetroot (1.8oz.) | | | | | | | | | |
| Coleslaw (1.8oz.) | | | | | | | | | |
| Avocado (1.8oz.) | | | | | | | | | |
| Baked beans (1.8oz.) | | | | | | | | | |
| Dried lentils, beans, peas (1.8oz.) | | | | | | | | | |
| Tofu, soya meat, TVP, Vegeburger (1.8oz.) | | | | | | | | | |

2. Are there any **OTHER** foods which you ate more than once a week? Yes ☐ No ☐

**If yes,** please list below

| Food | Usual serving size | Number of times eaten each week |
|---|---|---|

3. What type of milk did you most often use?

**Select one only**

- Full cream, silver ☐
- Skimmed/blue ☐
- Dried milk ☐
- Other, *specify* ☐
- Semi-skimmed, red/white ☐
- Channel Islands, gold ☐
- Soya ☐
- None ☐

4. How much milk did you drink each day, including milk with tea, coffee, cereals etc?

- None ☐
- Quarter of a pint ☐
- Half a pint ☐
- Three quarters of a pint ☐
- One pint ☐
- More than one pint ☐

5. Did you usually eat breakfast cereal (excluding porridge and Ready Brek mentioned earlier)?

Yes ☐ No ☐

**If yes,** which brand and type of breakfast cereal, including muesli, did you usually eat?

**List the one or two types most often used**

| Brand *e.g. Kellogg's* | Type *e.g. cornflakes* |
|---|---|

6. What kind of fat did you most often use for frying, roasting, grilling etc?

**Select one only**

- Butter ☐
- Lard/dripping ☐
- Vegetable oil ☐
- Solid vegetable fat ☐
- Margarine ☐
- None ☐

**If you used vegetable oil**, please give type eg. corn, sunflower ☐

7. What kind of fat did you most often use for baking cakes etc?

**Select one only**

- Butter ☐
- Lard/dripping ☐
- Vegetable oil ☐
- Solid vegetable fat ☐
- Margarine ☐
- None ☐

**If you used margarine**, please give name or type eg. Flora, Stork ☐

8. How often did you eat food that was fried at home?

Daily ☐ 1-3 times a week ☐ 4-6 times a week ☐
Less than once a week ☐ Never ☐

9. How often did you eat fried food away from home?

Daily ☐ 1-3 times a week ☐ 4-6 times a week ☐
Less than once a week ☐ Never ☐

10. What did you do with the visible fat on your meat?

Ate most of the fat ☐ Ate as little as possible ☐
Ate some of the fat ☐ Did not eat meat ☐

11. How often did you eat grilled or roast meat? ☐☐ times a week

12. How well cooked did you usually have grilled or roast meat?

Well done /dark brown ☐ Lightly cooked/rare ☐
Medium ☐ Did not eat meat ☐

13. How often did you add salt to food while cooking?

Always ☐ Rarely ☐
Usually ☐ Never ☐
Sometimes ☐

14. How often did you add salt to any food at the table?

Always ☐ Rarely ☐
Usually ☐ Never ☐
Sometimes ☐

15. Did you regularly use a salt substitute (eg LoSalt)? Yes ☐ No ☐

**If yes**, which brand? ______________________

16. During the course of last year, on average, how many times a week did you eat the following foods?

| **Food type** | **Times/week** | **Portion size** |
|---|---|---|
| Vegetables (not including potatoes) | ☐☐ | medium serving |
| Salads | ☐☐ | medium serving |
| Fruit and fruit products (not including fruit juice) | ☐☐ | medium serving or 1 fruit |
| Fish and fish products | ☐☐ | medium serving |
| Meat, meat products and meat dishes (including bacon, ham and chicken) | ☐☐ | medium serving |

17. Have you taken any vitamins, minerals, fish oils, fibre or other food supplements during the past year? Yes ☐ No ☐ Don't know ☐

**If yes,** please complete the table below. If you have taken more than 5 types of supplement please put the most frequently consumed brands first.

| **Vitamin supplements** | **Average frequency** Tick **one** box per line to show how often on average you consumed supplements | | | | | | | | | |
|---|---|---|---|---|---|---|---|---|---|---|
| **Name and brand** Please list full name, **brand** and strength | **Dose** Please state number of pills, capsules or teaspoons consumed | Never or less than once a month | 1-3 per month | Once a week | 2-4 per week | 5-6 per week | Once a day | 2-3 per day | 4-5 per day | 6+ per day |

*Thank you for your help*
