## Supplemental Appendix 3 for "A feasibility study to test a novel approach to dietary weight loss with a focus on assisting informed decision making in food selection"

**S3 Appendix. Exit survey outcomes**

| Category | Question | Mean | SD |
| --- | --- | --- | --- |
| Personal gain | 1. Do you feel as though you have learned valuable nutrition information? | 6.55 | 0.69 |
|  | 2. How effective was this program for achieving your objectives? | 5.73 | 1.35 |
|  | 3. Were the skills taught in this program applicable to your everyday life? | 6.18 | 0.60 |
| Mean |  | **6.15** | **0.97** |
| Program content | 4. Was the content valuable? | 6.55 | 0.69 |
|  | 5. Were the instructors knowledgeable? | 7.00 | 0.00 |
|  | 6. Did the instructors relay the information in a manner that was understandable for you? | 6.27 | 0.65 |
|  | 7. Was the set-up of the course (face-to-face sessions, individual sessions, e-mail communication) useful for you? | 6.18 | 0.75 |
|  | 8. Were the instructors readily available to offer you support? | 6.91 | 0.30 |
| Mean |  | **6.58** | **0.65** |
| Materials and feedback | 9. Were the presentations and feedback forms presented in a manner that was easy for you to understand? | 6.27 | 0.65 |
|  | 10. Was the feedback provided helpful? | 6.64 | 0.67 |
| Mean |  | **6.45** | **0.73** |
| Engagement | 11. How engaging was the course for you? | 5.64 | 1.03 |
|  | 12. Were there enough interactive opportunities for you (with the instructors and/or other participants)? | 6.27 | 0.65 |
| Mean |  | **5.95** | **0.88** |
| Overall evaluation | 13. Was the program worth your time? | 6.55 | 0.82 |
|  | 14. Would you recommend this program to others? | 6.64 | 0.92 |
| Mean |  | **6.59** | **0.85** |

Choices of responses are: “Definitely yes”, “Yes”, “A little bit”, “Neutral”, “Not really”, “No” and “Definitely not”. A score ranges from 1 to 7, with seven being “Definitely yes”. n=11.
