## Supplementary material for "A feasibility study to test a novel approach to dietary weight loss with a focus on assisting informed decision making in food selection": Study Protocol

### NEW PROTOCOL APPLICATION

Application for Review of Research Involving Human Subjects

| This Section is for Office Use Only | |
|---|---|
| University of Illinois IRB Protocol No. ______________________ | Track: ____________________ |
| Exempt under 45 CFR §46.101(b) ☐(1) ☐(2) ☐(3) ☐(4) ☐(5) ☐ (6) | Reviewer 1: ________________ |
| Expedite, Category ☐(1)☐(2)☐(3)☐(4)☐(5)☐(6)☐(7)☐(8)☐(9) | Reviewer 2: ________________ |

☐ Initial Submission, date of submission ____________

☐ Revised New Protocol Application, date of revised New Protocol Application ____________

**1. RESPONSIBLE PROJECT INVESTIGATOR (RPI)** The RPI must be a nonvisiting member of the University of Illinois faculty or staff who will serve as project supervisor at Illinois. **For other research team members [including those from other institutions], please complete the Research Team Attachment and provide with the completed application.** Include all persons who will be 1) directly responsible for the project's design or implementation, 2) recruitment, 3) obtain informed consent, 4) involved in data collection, data analysis, or follow-up.

| | | | |
|---|---|---|---|
| **Last Name:** Nakamura | First Name: Manabu | | Academic Degree(s):PhD |
| **Dept. or Unit:** Food Science and Human Nutrition | Office Address: 463 Bevier Hall | | Mail Code: 182 |
| **Street Address:** 905 S. Goodwin Ave | City: Urbana | State: Illinois | Zip Code: 61801 |
| **Phone: (217)-333-1267** | **Fax: 217-265-0925** | | |
| **Urbana-Champaign Campus Status: Nonvisiting member of (Mark One)**<br>☒**Faculty** ☐ **Academic Professional/Staff** | | | |
| **Training**<br>☒ **CITI Training, Date of Completion, 7/21/2017**<br>☐ **Additional training, Date of Completion[1],** | | | |

**2. PROJECT TITLE**

| |
|---|
| Individualized Diet Improvement Program (iDip) for weight loss and maintenance. |

**3. FUNDING** Indicate whether this research is funded by, or application has been made for, a grant, contract, or gift.

**3A. STATUs** ☐ Research is **not funded** and is **not pending** a funding decision (Proceed to Part 4).

☒ Research is **funded** (funding decision has been made).

☐Funding decision is **pending**. Funding proposal submission date.

**3B. SOURCE(S)** If the research is funded or pending a funding decision, mark and name all sources:

| **Type of Funding—check all that apply** | **Name of Source** |
|---|---|

[1] Additional CITI modules may be required depending on subject populations or types of research. These include: (i) research enrolling children; (ii) research enrolling prisoners; (iii) FDA regulated research; (iv) data collected via the internet; (v) research conducted in public elementary/secondary schools; and, (vi) researchers conducted in international sites

| | | |
|---|---|---|
| ☐ | **University of Illinois Department, College, or Campus** (includes Research Board and Campus Fellowship Training Grants) | |
| ☒ | **Federal** (from federal agencies, offices, departments, centers) | **USDA HATCH** |
| ☐ | **Commercial Sponsorship & Industry**[2,3] (from corporations, partnerships, proprietorships) | |
| ☐ | **State of Illinois Department or Agency** (from any state office or entity) | |
| ☐ | **Gift or Foundation (including UIF)** (public or private foundations, not-for-profit corporations, private gifts) | |

→ Check here if the funding is through a Training Grant: ☐

**3C. PROPOSAL** Attach a complete copy of the funding proposal or contract. ☒ Attached

Sponsor-assigned grant number, if known: **ILLU-698-908**

Title of Funding Proposal or Contract, if different from Project Title in Part 2:

| |
|---|
| Knowledge-based dietary weight management. |

**3D. FUNDING AGENCY OFFICIAL, IF ANY, TO BE NOTIFIED OF IRB APPROVAL**

| | | |
|---|---|---|
| Last Name: not applicable | First Name: | Salutation: |
| Agency: | Office Address: | Mail Code: |
| Street Address: | City: | State: / Zip Code: |
| Phone: | Fax: | E-mail: |

**4. FINANCIAL INTERESTS:** Indicate below if any investigators or any members of their immediate families have any relationships, commitments, or activities with the sponsor of this research that might present or appear to present a conflict of interest with regard to the outcome of the research. (If a financial conflict of interest exists, please submit the University of Illinois approved conflict management plan. If you have questions about conflict of interest contact the Office of the Vice Chancellor for Research at 217-333-0034.)

☐ Ownership, equity or stock options
☐ Has been disclosed to the Illinois campus **OR** ☐ has not been disclosed to the Illinois campus

☐ Personal compensation such as royalties, consulting fees etc.
☐ Has been disclosed to the Illinois campus **OR** ☐ has not been disclosed to the Illinois campus
☐ Intellectual property such as patents, trademarks, copyright, licensing, etc.
☐ Has been disclosed to the Illinois campus **OR** ☐ has not been disclosed to the Illinois campus

☐ Other conflict of interest:
☐ Has been disclosed to the Illinois campus **OR** ☐ has not been disclosed to the Illinois campus

☒ No conflicts exist

**5. SUMMARIZE THE RESEARCH.** In **LAY LANGUAGE**, summarize the objectives and significance of the research.

[2] Clarify whether or not sponsor requires specific language in the contractual agreement that impacts human subjects research [2]

[3] Clarify whether or not the sponsor requires the protocol adhere to ICH GCP (E6) standards

| |
|---|
| The aim of this research is to develop a weight loss and weight maintenance program through dietary modifications for adults with obesity. Although scientific studies have shown feasibility of rapid and safe dietary weight loss and subsequent weight maintenance, no efficacious dietary weight management program is widely available, and thus bariatric surgery remains as the most reliable approach for weight loss/management. Safe and effective dietary weight loss and subsequent weight maintenance requires flexible, individualized advice by an experienced dietitian/nutritionist. The specific objectives of this research project are to 1) develop a cost-effective weight loss and maintenance program and 2) provide a training opportunity for weight management instructors. |

**6. PERFORMANCE SITES**

| Including Urbana-Champaign sites, describe ALL the research sites for this protocol. For each non-Urbana-Champaign site, describe: Whether the site has an IRB. Whether the site has granted permission for the research to be conducted. Contact information for the site. If the site has an IRB, whether the site's IRB has approved the research or planned to defer review to a University of Illinois IRB. | | For non-Illinois sites, documentation of IRB approval is: |
|---|---|---|
| 1. | Department of Food Science and Human Nutrition, UIUC | ☐ Attached ☐ Will Follow ☐ N/A |
| 2. | | ☐ Attached ☐ Will Follow ☐ N/A |
| 3. | | ☐ Attached ☐ Will Follow ☐ N/A |

List and describe any additional Performance Sites information on an attachment and check here: ☐

**7. DESCRIBE THE HUMAN SUBJECTS**

**7A. SECONDARY DATA ONLY?** If this research *only* involves the analysis of data that *has already been collected* from human subjects and *no new data collection will occur,* check here: ☐

**7B. MATERIALS OF HUMAN ORIGIN?** Will this research involve the collection, analysis, or banking of human biological materials (*e.g.,* cells, tissues, fluids, DNA)? ☐ **Yes** ☐ **No**
If yes attach **Appendix C**, the *Biological Materials Form.*

**7C. ANTICIPATED NUMBERS** How many subjects, including controls, will you study in order to get the data that you need? If you plan to study disproportionate numbers of a given sex, race, or minority group, provide scientific rationale in Part 11.

| **Performance Site** | | **# Male** | **# Female** | **Total** |
|---|---|---|---|---|
| 1. | Department of Food Science and Human Nutrition, UIUC | 90 | 90 | 180 |
| 2. | | | | |
| 3. | | | | |
| **TOTALS** | | **90** | 90 | 180 |

List Anticipated Numbers for additional Performance Sites on an attachment and check here: ☐

**7D. AGE RANGE** Mark all that apply. Researchers planning to include children in research projects involving *more than minimal risk* must provide written documentation of the benefits that are likely to accrue to a child participating in the project. This should include information gathered on adults, if it exists, or an explanation about why it does not exist.

☐ 0–7 years ☐ 8–17 years ☒ 18–64 years ☐ 65+ years

→ ☐ If applicable, written documentation of benefits for including children in ***more than minimal risk*** research is attached.

**7E. SPECIAL OR VULNERABLE POPULATIONS** Mark groups that will be targeted by design. Also indicate groups likely to be involved in the research even though they are not targeted by design.

☒ None of the following special populations will be targeted

| | |
|---|---|
| ☐ Children (age < 18 years) | |
| ☐ Neonates | ☐ Mentally disabled or cognitively impaired persons |
| ☐ Fetuses *(in utero)* | ☐ Adults with legal guardians |
| ☐ *in vitro* fertilization subjects | ☐ Persons with limited civil freedom (*e.g.,* prisoners) |
| ☐ Pregnant or lactating women | ☐ Specific racial or ethnic group(s)— describe: |
| ☐ Inpatients | ☐ Low income or economically disadvantaged persons |
| ☐ Outpatients | ☐ Illinois Students—name subject pool, if applicable: |
| ☐ Elderly (age > 65 years) | ☐ Other College Students—name subject pool, if applicable: |
| ☐ Other (describe here): | |

**7F.** if you checked any of the groups in question 7E, describe additional safeguards included in the protocol to protect the rights and welfare of special or vulnerable populations.

**8. RECRUITMENT**

**8A-1 RECRUITING PROCEDURES** specifically describe the systematic procedures for finding and recruiting subjects or requesting pre-existing data or materials. 1) State whether any of the researchers are associated with the subjects (*e.g.,* subjects are students, employees, patients). 2) Name any specific agencies or institutions that will provide access to subjects or subject data. 3) Who will contact the prospective subjects? 4) Who gives approval if subjects are chosen from records? 5) Describe solicitation through the use of advertising (*e.g.,* posters, flyers, announcements, newspaper, radio, television, Internet), face-to-face interaction, direct mail or phone contact, classrooms, subject pools, health care registries, patient referrals, and institutional "gatekeepers," as applicable.

The participants will be recruited from Urbana-Champaign, Illinois, and surrounding areas by electronic bulletin boards, flyers and word of mouth. Interested individuals will contact the investigators via electronic mail to obtain detailed information of the study. After they read the general description of the study, individuals who want to be in the study will send an email to the investigators indicating their interest. Then, the investigators will send the Consent Form and the Medical History Form to the participants. They will fill the forms and send them back to investigators via mail to 463 Bevier Hall, 905 S. Goodwin Avenue, Urbana, IL 61801. The eligible participants will be notified via email and schedule an individual meeting. Written informed consent signed by both parties will be obtained at or within a week from this individual session.

1. The researchers will not be directly associated with the participants; however, some participants may be employees within the Department of Food Science and Human Nutrition.
2. No specific agencies or institutions will provide access to the subjects or subject data.
3. Prospective subjects will contact investigators via electronic mail to obtain information of the study.
4. Subjects will not be chosen from records; therefore, no approval is needed.

5. Prospective participants will be recruited through university electronic-mail notices and flyers posted in local communities, including the University of Illinois, Urbana-Champaign, campus and surrounding areas.

**8 A-2 Attach final copies of recruiting materials** including the final copy of printed advertisements and the final version of any audio/taped advertisements and check here:
Attached ☒ Will Follow ☐

**8B. WITHHELD INFORMATION** Do you propose to withhold information from subjects prior to or during their participation?
☐ Yes ☒ No

If yes, describe what will be withheld, justify the withholding (address risks, provide rationale), describe the debriefing plan, and attach a labeled copy of a written debriefing form, to be provided to subjects. ☐ Debriefing Attached ☐ Will Follow

**8C. PROTECTED HEALTH INFORMATION (PHI)** The IRB must address the privacy and use of health information that is created, received, or housed by health care providers, health plans, or health care clearinghouses and that identifies or could be used to identify an individual. During *either recruiting or data collection*, will you use or have access to such information that is related to the past, present or future health or conditions of a *living or deceased* individual, provision of health care to the individual, or the payment for the provision of health care to the individual? ☒ Yes ☐ No

**8D. SCHOOL-BASED RESEARCH** If subjects will be recruited from Illinois public or private elementary or secondary schools, additional deadlines and procedures apply. Criminal background clearances might be required. Special consideration must be given to the exclusion of protected populations. Please contact the Office of School–University Research Relations (OSURR) (217.244.0515 or http://www.ed.uiuc.edu/BER/OSURR.html) for more information. Mark one:

☐ Illinois schools **will** be used ☒ Illinois schools **will not** be used

**9. INCLUSION AND EXCLUSION CRITERIA** Address all four of the following items in explaining who will and will not qualify for participation and how that determination will be made: (1) Describe procedures to assure equitable selection of subjects. Justify the use of any special or vulnerable groups marked in Part 7E. Selection criteria that target one sex, race, or ethnic group require a clear scientific rationale. (2) List specific criteria for inclusion and exclusion of subjects in the study, including treatment groups and controls. (3) Name and attach copies of measures and protocols that will be used to screen applicants. (4) Explain how the inclusion/exclusion criteria will be assessed and by whom. If special expertise is required to evaluate screening responses or data, tell who will make this evaluation and describe their training and experience.

1. We will assure equitable selection of subjects by enrolling participants from the eligible applicants to reflect demographics of the local community.

2. Inclusion criteria will include:
-age 18-64 years;
-body mass index (BMI) >28 $kg/m^2$;
-not currently pregnant or lactating;
-have Wi-Fi at home, a working email, and a smartphone.
-no self-reported severe metabolic, cardiovascular or musculoskeletal disease;
-not using insulin injection;
-able to attend the 22 (1 hour) diet improvement sessions.

-willing to lose 20 lb. or more during 6 months and maintain a healthy weight for one year.
-keep weighing their weight for one and a half year.
-fluent in reading and writing English.

Exclusion criteria will include:
-age <18 or >64 years;
-body mass index of <28 kg/m$^2$;
-currently pregnant;
-currently lactating;
-self-reported with sever metabolic, cardiovascular or musculoskeletal disease;
-use insulin injection;
-not able to attend the 22 (1 hour) diet improvement sessions for 1 year;
-failed to set up Wi-Fi scale;
-failed to submit food frequency questionnaire

3. Measures and protocols used to screen applicants will include:
-Medical History Form (included with screening forms);
-Food Frequency Questionnaire (FFQ, included with screening forms);
-Anthropometric measurement at an individual meeting to confirm BMI > 28

4. Prospective participants will self-assess eligibility based on inclusion/exclusion criteria sent to them. Upon return of screening materials and forms, participants will be assessed for full eligibility by the study investigators. No special training or expertise is required to evaluate screening responses and/or data. Regarding all forms and documents that ineligibility participants have provided to the investigators will be destroyed immediately after they have been notified of ineligibility.

**10. RESEARCH PROCEDURES: Using LAYMAN'S LANGUAGE,** specifically describe what the participants (treatment groups and controls) will do and where the research activities will take place. Give approximate dates and durations for specific activities, including the total number of treatments, visits, or meetings required and the total time commitment. (For schools-based research where class time is used, describe in detail the activities planned for nonparticipants and explain where (*e.g.,* in a classroom, in a private area) both participants and nonparticipants will be located during the research activities. Include a concise description of procedures, locations, time commitments, and alternate activities on the relevant consent and assent forms.)

Screening: Interested individuals will contact investigators via electronic mail to obtain a general description of the study. The participants who want to be in the study will send an email indicating their continued interest after they read the general description of the study. Then, the investigators will send the Consent Form without the signature line and the Medical History Form to the participants, and they will read the consent form carefully without signing it out and fill out the medical history form and send the forms to investigators via mail to 463 Bevier Hall, 905 S. Goodwin Avenue, Urbana, IL 61801. The sealed envelope will not be provided so; It will cost approximately 1.50 -2 $ each participant. Once the investigators receive the forms, they will analyze and review the forms. Eligible participants will be informed via email to schedule an individual meeting appointment. The participants, who are on the waiting list, will be notified and set up an individual meeting in future when the study will be expanded. Regarding the ineligible participants, all their forms will be destroyed immediately after they have been informed of ineligibility.

Individual meeting: At the Individual meeting, to be held in Bevier Hall on the University of Illinois, Urbana-Champaign campus, the investigators will explain the purpose, requirements, expectations, risks and benefits of the 1.5-year study. Also, the investigators will take body measurements, including: height, weight, waist, hip circumference, and blood pressure. The investigators will review the Informed Consent Form and ask the participants to fill out the food frequency questionnaire (FFQ). Also, the investigators will setup the Wi-Fi scale for each participant and address any questions or concerns expressed by participants. This session will last approximately 1 hour. All participants will receive a schedule of meeting dates. Enrolled participants will return their signed Informed Consent Forms in the individual meeting.

Eligible participants will then be in an intervention group (n ~ 30) led by a registered dietitian, or the wait-list group (n ~ 30). The wait-list group will be enrolled in the program 6 months later. This enrollment pattern will be repeated every 6 months for 3 years. Each participant will be assigned a 3-digit ID code number. Additional anthropometry sessions will be scheduled for each participant at months 6, 12 and 18 months.

Intervention groups: During a 12-month period, participants will attend total of 22 diet improvement sessions, each of which will last approximately 1 hour. Twelve sessions will be held in the first 5 months, and will be focused on safe and efficient weight loss. The participants will learn to create personalized weight loss diet from their kitchen based on their diet practice and food preference. The next 10 sessions will be held in the last 7 months, and will be focused on weight maintenance and healthy eating. The participants will build skills to select foods and create meals that prevent them from overeating.

Data collection: The daily body weight will be automatically collected using a Wi-Fi scale. Anthropometric and blood pressure measurements will be collected at baseline (week 0), 6, 12 and 18 months at individual meetings, which will take 15 minutes to 1 hour each. In addition to FFQ at the beginning, participants will complete one or more 24-hour food records at home, and bring these to the next session for diet analysis as needed.

Safety and efficacy measure of dietary intervention:
The efficacy of the program will be constantly monitored by the daily body weight data obtained via Wi-Fi scale. Safety of the participants during weight loss will be maintained by daily weight monitoring, 24-hour food records and individual advising.

Dietary intake assessment:
Participants will be taught to accurately complete FFQ to have a clear picture about the dietary habits of the participants. The investigators will analyze the FFQ and provide the participants with individualized advice and options for improvement of diet. Then, the participants will make a decision for changing their diet. The FFQ will be administered 3 times during the study at 0, 6, and12 month to monitor the diet changes of participants.

Also, the participants will be taught to accurately complete 24-hour food records using a food record form and handouts that contain examples of standard serving sizes of different foods and beverages. Investigators use the 24-hour record to check adequacy of their diet and to troubleshoot if a participant is not losing weight effectively. Participants will receive an individualized advice based on 24-hour food record analysis.

Anthropometric and blood pressure measurements:
Standing height (cm) will be measured using a calibrated scale-mounted stadiometer (Seca 700, Hanover, MD) to the nearest 0.1 cm. Body weight will be measured on a calibrated balance beam scale (Seca 700, Hanover, MD) to the nearest 0.1 kg. Body mass index ($kg/m^2$) will be calculated based on height and body weight measurements. Waist and hip circumference (cm) will be measured and recorded to the nearest 0.1 cm using a retractable tape measure (Gulik II, Gay Mills, WI) and will be used to calculate waist:hip ratio.

Seated systolic and diastolic blood pressure (mm Hg) will be measured by a standard sphygmomanometer (Baumanometer® Desk Model, Copiague, NY) after a 5-min rest period. Two measurements will be taken with a 2- to 3-min rest period between readings. The two systolic arterial pressure values will be averaged, and the two diastolic arterial pressure values will be averaged; these means will be used in data analyses. Measurements will be completed by a trained study investigator, who will be blinded to the intervention assignment.

Daily body weight will be automatically collected using a Weight Gurus Wi-Fi Smart Scale model No. 0385 (Greater Goods, St Louis, MO). The investigators have access to each participant to tracking their daily weight changes throughout the weight gurus website.

Participants are to be available for the full duration of this 18-month intervention; however, participants are permitted to take vacations and continue their schedules as usual. Absences and vacations will be expected and accommodated with attention given to scheduling of data collection sessions as well as educational sessions in order to minimize participant burden. Make-up sessions will be offered and educational materials will be mailed to participants if necessary.

**11. EQUIPMENT** Will any physical stimulation or physiological data acquisition equipment be used with the subjects?
☒ Yes ☐ No If yes, attach **Appendix A**, the *Research Equipment Form.*

**12. DEVICES** Will any devices be used with the subjects?
☐ Yes ☒ No If yes, attach **Appendix B-1,** the *Device Form*.

**13. DRUGS AND BIOLOGICS** Will any drugs or chemical or biological agents be used with the subjects?

☐ Yes ☒ No    If yes, attach **Appendix B-2**, the *Drug and Chemical Usage Form*.

**14. MRI AT BIC** To use the Beckman Institute Biomedical Imaging Center (BIC) in human subject's research, you must obtain *prior approval* from the BIC (217.244.0600;) and use BIC-approved screening and consent forms. Attach:

BIC approval ☐ Attached
BIC screening form ☐ Attached
BIC consent form ☐ Attached

**15. MEASURES** If subjects will complete questionnaires, surveys, interviews, psychological measures, or other measures, however administered, the IRB must review and approve the measures. List all such measures here and attach complete, labeled copies (including translations, if applicable) to this application:

| | | |
|---|---|---|
| Measure 1: | Medical History Form (screening) | ☒ Attached ☐ Will Follow |
| Measure 2: | Food Frequency Questioners (individual meeting) | ☒ Attached ☐ Will Follow |
| Measure 3: | 24-hr Food Record ( diet improvement sessions) | ☒ Attached ☐ Will Follow |
| Measure 4: | | ☐ Attached ☐ Will Follow |

List additional Measures on an attachment and check here: ☐

**16. SUBJECT REMUNERATION**

Will subjects receive inducements or rewards before, during, or after participation?
☐ Yes ☒ No
If yes, will payment be prorated for partial participation?
☐ Yes ☐ No
If remuneration will be given, for each subject group:
(1) Specify the form of remuneration, including $, course credit, lottery, gift certificate, or other;
(2) State the $ amount or the approximate $US value, or the course credit and its percentage of the final grade;
(3) Explain the remuneration plan, including whether and how prorating will be made for partial participation;
(4) For lotteries, include (a) the number of prizes, (b) the nature and value of each prize, (c) the approximate odds of winning, (d) the date(s) of the drawing(s), and (e) how winners will be notified, by whom, and by when; and
(5) Include all this information on the relevant consent forms.

Participants will be allowed to keep a Wi-Fi Scale when they complete the one and half year of the study; however, the participants must return the scale if they are unable to complete the study. There will be no monetary compensations.

**17. SUBJECT OUTLAY** Will subjects incur costs for research-related procedures (*e.g.*, longer hospitalization, extra tests), use of equipment, lost compensation, or transportation (over 50 miles)? ☐ Yes ☒ No    If yes, describe here:

**18. CONFIDENTIALITY OF DATA** Answer each of the following to describe methods that will ensure the confidentiality of individually identifiable data. Confidentiality is required unless subjects give express, written permission to have their identifiable information published, presented, or shared.

**18A. CHECK IF USED IN DATA COLLECTION:** ☐ Audio tapes/digital voice ☐ Video tapes ☐ Still photos ☐ Other imaging

**18B. DATA COLLECTION** Explain how the data will be collected. If anonymous data collection is proposed, provide details of how investigators *will not have the ability to trace responses to subject identities.* For multiphase data collection or if multiple contacts will be made with subjects, specifically explain the subject tracking and coding systems.

Address the confidentiality of data collected via e-mail, databases, Web interfaces, computer servers, and other networked information, as applicable.

Confidentiality of data collected will be maintained by assigning each participant a 3-digit ID code which will be used to enter all data in an electronic database. ID codes will be used on all documents and materials as the only form of identification; no names will be used. All files and participant documentation will be stored in locked filing cabinets, and only the graduate researcher will have access to the key. Electronic documents, forms and data will be stored on a password protected computer. Only the primary investigator, co-investigator, and research team will have access to stored data. Any data collected via email will be completed using ID codes and deleted and emptied from trash following data collection. Only the primary investigator will have access to the ID code sheet.

**18C. DATA SECURITY** Describe how and where the data be kept so that the data remain confidential.

All hard copies of completed forms, documents and other study materials will be kept in locked filing cabinets in the laboratory space. Electronic data will be stored on a password protected computer. Only the principal investigator, co-investigator and research team will have access to stored data.

**18D. STAFF TRAINING** Describe the training and experience of all persons who will collect or have access to the data.

Nakamura, Manabu, PhD, is an Associate Professor in Department of Food Science and Human Nutrition (FSHN), and will serve as the Responsible Project Investigator and will be responsible for overseeing the development, coordination and administration of the project. He will supervise graduate students and will oversee participant recruitment, screening and intervention sessions. Additionally, he will supervise other individuals of the research team. He will facilitate and aid in preparation of scientific abstracts, manuscripts, presentations and progress reports. His area of expertise is biochemistry and physiology of adaptive response of our body to weight gain and weight loss.

Applegate, Catherine C, MS, RDN, is a graduate research assistant and will primarily facilitate curriculum development under the supervision of the principal investigator, will teach the group sessions, and provide individual counseling throughout the program. She will serve as the in-person research representative and educator during each group and individual session and will discuss progress reports with individuals. She has undergone an accredited didactics internship program and has received the certificate for training of weight management through the Commission on Dietetic Registration in addition to her completed Masters in Human Nutrition degree and current progress towards a doctoral degree in Nutritional Sciences.

Emamaddin, Abrar, BS, is a graduate student in the department of Food Science & Human Nutrition and will have primary responsibilities for participant recruitment, screening, enrollment and retention. She will serve as the primary point of contact for participants and will be in charge of scheduling all appointments, data collection sessions and educational sessions. She will administer and manage study forms, documents and materials, including dietary intake questionnaires, for the duration of the trial. She will participate in all data collection sessions and management of data. She will be involved in the preparation and submission of progress reports. Her research interests include weight loss and maintenance across the weight management spectrum. She has served as the lead investigator for a qualitative study on weight-loss maintenance and has participated in data analyses and preparation of manuscripts. She completed her undergraduate degree in clinical nutrition.

Additional staff trained in laboratory procedures and data management and analyses will be used as necessary. Once these individuals have been identified, an IRB amendment will be submitted adding these project staff members.

All unidentified staff members will complete CITI training before participation in any component of this research.

**18E. DATA RETENTION** How long will the data be kept?

| |
|---|
| Data will be maintained for three years after completion of the study |

**18F. DISSEMINATION OF RESULTS** what is (are) the proposed form(s) of dissemination (*e.g.,* journal article, thesis or academic paper, conference presentation, sharing within industry or profession)?

| |
|---|
| Findings will be disseminated via abstracts and presentations at scientific conferences and through manuscripts and journal articles within the field. |

**18G. PRIVACY** Describe provisions to protect the privacy interests of subjects.

| |
|---|
| Confidentiality of data collected will be maintained by assigning each participant a three-digit identification (ID) code which will be used to enter all data in an electronic database. ID codes will be used on all documents and materials as the only form of identification; no names will be used. All files and participant documentation will be stored in locked filing cabinets. Electronic documents, forms and data will be stored on a password protected computer. Only the primary investigator, co-investigator, and research team will have access to stored data. Any data collected via email will be completed using ID codes and deleted and emptied from trash following data collection. The primary investigator will immediately destroy documents that are obtained from individuals who do not qualify for the study, as determined during the pre-screening and screening steps. These documents will be shredded. |

**18H. INDIVIDUALLY IDENTIFIABLE INFORMATION** Will any individually identifiable information, including images of subjects, be published, shared, or otherwise disseminated? ☐ Yes ☒ No

**If yes,** subjects must provide explicit consent or assent for such dissemination. Provide appropriate options on the relevant consent documents.

**19. INFORMED CONSENT:** University policy requires the execution of a comprehensive, written document that is signed by the subject (or the subject's authorized representative) as the principal method for obtaining consent from subjects. The language in the document must be understandable to the subject or the subject's legally authorized representative.

An investigator may request a Waiver or Alteration of Informed Consent or a Waiver of Documentation of Informed Consent (e.g., online consent, oral consent). If requesting a waiver please complete the appropriate waiver form at: www.irb.illinois.edu and submit it with the New Protocol Application form for review.

**Children must *assent*** (or, voluntarily agree) to participation and a parent must separately consent on behalf of their child (*i.e.,* two different forms are generally required). Children under age 8 may assent either orally or passively, depending on their level of maturity. Children 8–17 years old should sign a written form unless the University of Illinois at Urbana-Champaign IRB approves a different process.

**19A. TYPE OF CONSENT** Check all that apply and attach one copy of each relevant form, letter, or script on university letterhead. Include translations, if consent will be obtained in a foreign language. Use headings, headers, or footers to uniquely identify each document and associate it with the subject group for which it will be used.

☒ **Written informed consent (assent) with a document signed by**
☒ adult subjects ☐ parent(s) or guardian(s) ☐ adolescents aged 8–17 years

☐ **Waiver or Alteration of Informed Consent (Attach waiver form.)**
☐ adult subjects ☐ parent(s) or guardian(s) ☐ adolescents aged 8–17 years

☐ **Waiver of Documentation (signature) of Informed Consent (Attach waiver form.)**
☐ adult subjects ☐ parent(s) or guardian(s) ☐ adolescents aged 8–17 years

**19B. USE OF PROXY** Will others (*e.g.,* next of kin, legal guardians, powers of attorney) act on behalf of adult subjects in giving consent to participate in this research? ☐ Yes ☒ No (if yes, describe in Section 20D.)

**19C. USE OF PROXY OUTSIDE THE UNITED STATES** If a proxy is used in research conducted outside Illinois, provide justification (e.g., statement of an attorney or copy of applicable law) that the proxy is authorized under the laws of the jurisdiction in which the research will be conducted to consent to the procedures involved in this protocol.

| |
|---|
| Not applicable. |

**19D. CONSENT PROCESS** Describe when and where voluntary consent will be obtained, how often, by whom, and from whom. If cognitively impaired subjects (including children under age 8) will be involved, explain how the subject's understanding will be assessed and how often; include the questions that will be asked or actions that will be taken to assess understanding.

Describe any waiting period between informing the prospective subject and obtaining the consent. Describe steps taken to minimize the possibility of coercion or undue influence. Indicate the language used by those obtaining consent. Indicate the language understood by the prospective subject or the legally authorized representative.

If the research involves pregnant women, fetuses, or neonates, indicate whether consent will be obtained from the mother, father, or both. If the research involves children, indicate whether consent will be obtained from: Both parents unless one parent is deceased, unknown, incompetent, or not reasonably available, or when only one parent has legal responsibility for the care and custody of the child; or from one parent regardless of the status of the other parent.

| |
|---|
| Interested individuals will contact with investigators via electronic mail to obtain a general description of the study. The investigators will send a general description form to them. Then, the individuals who want to be in the study will send an email after they read the general description of the study. Then, the investigators will send the Consent Form and the Medical History Form to them and the participants will fill it out and send it to investigators in closed envelop via mail to 463 Bevier Hall, 905 S. Goodwin Avenue, Urbana, IL 61801. Once the investigators receive the completed forms, they will review the forms. The eligible participants will be noticed via email and schedule an individual meeting appointment. Written informed consent will be obtained during or within a week of the individual meeting. |

**20. RISKS**

**20A. DESCRIPTION** specifically describe all known risks to the subjects for the activities proposed and describe the steps that will be taken to minimize the risks. Include any risks to the subject's physical well-being, privacy, dignity, self-respect, psyche, emotions, reputation, employability, and criminal and legal status. Risks must be described on consent forms.

There are 2 potential risks: 1) nutrient deficiency during weight loss and 2) exposure of personal information.

Weight loss, if done correctly, is safe and does not affect health or daily physical and mental performance. However, incorrect weight loss attempt will cause nutrient deficiency and increase hunger and stress. This could happen if the participants choose an incorrect diet or provide us inaccurate information about their diet. The investigators will minimize the risk by close monitoring of daily weight changes, regular contact and administration of 24-dietary recall when necessary.

Investigators will keep all personal information shared during group sessions private, but it cannot be guaranteed that other participants will do the same. All participants will be informed about this risk and will be advised to decide carefully what information to share with the group.

There are no additional risks to participating in this study beyond those experienced in everyday life.

**20B. RISK LEVEL:** ☒ **No more than minimal risk**
(The probability and magnitude of harm or discomfort anticipated for participation in the proposed research are not greater in and of themselves than those ordinarily encountered in daily life or during the performance of routine physical or psychological examinations or tests).

☐ **More than minimal risk**

**20C. Data Monitoring Plan:** If you checked that the research is more than minimal risk, describe the provisions for monitoring the data to ensure the safety of subjects (Who will periodically monitor harms and benefits experienced by subjects to ensure that the relationship of risks to potential benefits remains unchanged? How often will monitoring occur? What analyses will be performed? If appropriate, what criteria will be used to stop the research based on monitoring of the results?)

**21. BENEFITS** Describe the expected benefits of the research to the subjects and/or to society.

| |
|---|
| The expected benefits to the participants are 1) regression of their health problems originated from overweight, and 2) building up knowledge and skills to create meals to lose weight and keep it off for the rest of their life. The expected benefits to the society by developing cost effective treatment of obesity are 1) reduction of morbidity and mortality caused by obesity, and 2) reduction of medical costs for treating complications of obesity. |

**22. RISK/BENEFIT ASSESSMENT** Weigh the risks with regard to the benefits. Provide evidence that benefits outweigh risks.

| |
|---|
| The risk involved by participating in the current study is minimal; as long as participants follow the guidelines set forth by the program objectives, there are only the benefits that accompany weight loss. |

If additional Risk/Benefit information is attached, check here: ☐

**23.** Is this a multi-center study in which the Illinois investigator is the lead investigator of a multicenter study, or the University of Illinois at Urbana-Champaign is the lead site in a multi-center study.

**Yes** ☐ **No**☒

If yes, describe the management and communication of information obtained that might be relevant to the protection of subjects, such as: unanticipated problems involving risks to subjects or others, interim results and protocol modifications.

| |
|---|
| Not applicable. |

**24. INVESTIGATOR ASSURANCES: The signature of the Responsible Project Investigator is required** (scanned or faxed signatures are acceptable). Other investigators are also responsible for these assurances and are encouraged to sign.

I certify that the information provided in this application, and in all attachments, is complete and correct.

I understand that I have ultimate responsibility for the protection of the rights and welfare of human subjects, the conduct of this study, and the ethical performance of this project.

I agree to comply with all Illinois policies and procedures, the terms of its Federal Wide Assurance, and all applicable federal, state, and local laws regarding the protection of human subjects in research.

I certify that

- The project will be performed by qualified personnel according to the University of Illinois at Urbana-Champaign IRB-approved protocol.
- The equipment, facilities, and procedures to be used in this research meet recognized standards for safety.
- No change will be made to the human subjects protocol or consent form(s) until approved by the University of Illinois at Urbana-Champaign IRB.
- Legally effective informed consent or assent will be obtained from human subjects as required.
- Unanticipated problems, adverse events, and new information that may affect the risk–benefit assessment for this research will be reported to the University of Illinois at Urbana-Champaign IRB Office (217.333.2670;) and to my Departmental Executive Officer.
- I am familiar with the latest information concerning IRB regulations and policies available at www.irb.illinois.edu, and I will adhere to the policies and procedures explained therein.
- Student and guest investigators on this project are knowledgeable about the regulations and policies governing this research.
- I agree to meet with the investigator(s), if different from myself, on a regular basis to monitor study progress.
- If I will be unavailable, as when on sabbatical or other leave, including vacation, I will arrange for an alternate faculty sponsor to assume responsibility during my absence. I will advise the Illinois IRB by letter of such arrangements.

I further certify that the proposed research has not yet been done, is not currently underway, and will not begin until IRB approval has been obtained.

| | | | |
|---|---|---|---|
| Responsible Principal Investigator | Date | Investigator | Date |
| Investigator | Date | Investigator | Date |

**25. (OPTIONAL) DEPARTMENTAL ASSURANCE** To be completed by the RPI's Departmental Executive Officer or their designee.

The activity described herein is in conformity with the standards set by our department and I assure that the principal investigator has met all departmental requirements for review and approval of this research.

Departmental Executive Officer (or designee) Date

***** For units that conduct **scientific merit review,** the signature above documents the following:

☐ 1. The research uses procedures consistent with sound research design.
☐ 2. The research design is sound enough to yield the expected knowledge.
